# Genomic dissection of the bacterial population underlying *Klebsiella pneumoniae* infections in hospital patients: insights into an opportunistic pathogen

**DOI:** 10.1101/2021.12.02.21267161

**Authors:** Claire L. Gorrie, Mirjana Mirceta, Ryan R. Wick, Louise M. Judd, Margaret M. C. Lam, Ryota Gomi, Iain J. Abbott, Nicholas R. Thomson, Richard A. Strugnell, Nigel F. Pratt, Jill S. Garlick, Kerrie M. Watson, Peter C. Hunter, David V. Pilcher, Steve A. McGloughlin, Denis W. Spelman, Kelly L. Wyres, Adam W. J. Jenney, Kathryn E. Holt

**Author notes:** Corresponding authors: CLG, KEH.

## Abstract

*Klebsiella pneumoniae* is a major cause of opportunistic healthcare-associated infections, which are increasingly complicated by the presence of extended-spectrum beta-lactamases (ESBLs) and carbapenem resistance. We conducted a year-long prospective surveillance study of *K. pneumoniae* clinical isolates identified in a hospital microbiological diagnostic laboratory. Disease burden was two-thirds urinary tract infections (UTI; associated with female sex and age), followed by pneumonia (15%), wound (10%) and disseminated infections/sepsis (10%). Whole-genome sequencing (WGS) revealed a diverse pathogen population, including other species within the *K. pneumoniae* complex (18%). Several infections were caused by *K. variicola/K. pneumoniae* species hybrids, one of which showed evidence of nosocomial transmission, indicating fitness to transmit and cause disease despite a lack of acquired antimicrobial resistance (AMR). A wide range of AMR phenotypes were observed and, in most cases, corresponding mechanisms were identified in the genomes, mainly in the form of plasmid-borne genes. ESBLs were correlated with presence of other acquired AMR genes (median 10). Bacterial genomic features associated with nosocomial onset of disease were ESBL genes (OR 2.34, p=0.015) and rhamnose-positive capsules (OR 3.12, p<0.001). Virulence plasmid-encoded features (aerobactin, hypermucoidy) were rare (<3%), and mostly present in community-onset cases. WGS-confirmed nosocomial transmission was rare (10% of cases) but strongly associated with ESBLs (OR 21, p<1×10^-11^). We estimate 28% risk of onward nosocomial transmission for ESBL-positive strains vs 1.7% for ESBL-negative strains. These data indicate the underlying burden of *K. pneumoniae* disease in hospitalised patients is due largely to opportunistic infections with diverse strains. However, we also identified several successful lineages that were overrepresented but not due to nosocomial transmission. These lineages were associated with ESBL, yersiniabactin, mannose+ K loci and rhamnose- K loci; most are also common in public clinical genome collections, suggesting enhanced propensity for colonisation and spread in the human population.

## Introduction

Healthcare-associated infections (HAIs) are common throughout the world, and in industrialised countries the burden of HAIs exceeds that of all other communicable diseases combined^1^. The major causative agents are opportunistic bacterial pathogens, which are generally viewed as commensals but can take advantage of the weakened immune system and altered microbiome of hospitalized patients to cause disease^2–5^. Particularly concerning are HAIs caused by Gram negative bacteria including *Klebsiella pneumoniae*, which is intrinsically resistant to ampicillin and increasingly displays acquired resistance to multiple additional drugs (multidrug resistance, MDR). Of particular concern, this organism readily acquires extended-spectrum beta-lactamase (ESBL) or carbapenemase genes that confer resistance to third-generation cephalosporins or carbapenems, respectively, leaving very limited options for antimicrobial therapy^6^. *K. pneumoniae* is amongst the leading causes of HAIs in hospitals globally, including urinary tract infections (UTI), pneumonia, wound infections and sepsis^2,7^. Recent studies confirm it is also a leading cause of neonatal sepsis in Africa^8^ and Asia^9^. *K. pneumoniae* is a clear opportunist, colonizing the human gut, nasopharynx and skin at high frequency, with food being a likely source of frequent exposure. *K. pneumoniae* is the ‘K’ in the ESKAPE pathogens, the group of opportunistic pathogens that together account for the majority of clinically significant MDR HAIs^3,10^, and ESBL and carbapenemase-producing (CP) *K. pneumoniae* combined make up the fastest-growing cause of drug-resistant infections in European hospitals^11^.

Asymptomatic *K. pneumoniae* colonization has been shown to be a source of HAIs, with attack rates estimated between 4-35% in colonized hospital inpatients^12–17^. Comparatively little is understood about *K. pneumoniae* pathogenesis, in contrast with closely related ‘true’ pathogens of the family Enterobacteriaceae, such as *Salmonella* or *Shigella*. Nearly all *K. pneumoniae* express the basic traits required for human infection, which are core to all strains: expression of O antigen lipopolysaccharide (LPS) and polysaccharide capsule (K antigen) (encoded by diverse K and O loci), the siderophore enterobactin (*ent* locus), and type I and type III fimbriae (*fim* and *mrk* loci^18,19^. So-called ‘hypervirulent’ *K. pneumoniae* clones – which are typically hypermucoid, aerobactin-producing, and carry the *K. pneumoniae* virulence plasmid (Kp-VP) and serum-resistant capsular (K) types such as K1, K2, K5^18,20,21^ – are associated with community-acquired invasive infections such as liver abscess and ophthalmitis^22,23^. However, there is no evidence such strains are more likely than others to cause opportunistic infections in the hospital setting. The acquired siderophore yersiniabactin (*ybt* locus) has been identified as a virulence factor relevant to nosocomial infection^22,24–26^; indeed a recent study of infection risk amongst patients colonized with CP sequence type (ST) 258 *K. pneumoniae* confirmed carriage of yersiniabactin (*ybt*) and modified O antigen synthesis as independent bacterial risk factors for subsequent infection^26^. *K. pneumoniae* also show propensity for nosocomial spread within the hospital environment, with sinks, drains, medical devices and cleaning products all being demonstrated as potential reservoirs of infection^27–29^. However, the relative contribution of patients’ own gut bacteria, vs nosocomial acquired bacteria, to the burden of *K. pneumoniae* HAI remains unclear, as does the question of whether bacterial features contribute to propensity for nosocomial infection.

Here we aimed to dissect the burden of *K. pneumoniae* HAIs in a tertiary hospital in Australia, via whole genome sequencing (WGS) of all clinical isolates identified in the hospital microbiological diagnostic laboratory for a one-year period. WGS studies have previously unveiled extensive diversity in the global *K. pneumoniae* population, which comprises hundreds of phylogenetically distinct lineages with variable gene content^18,22^. Additional diversity is harboured by related species in the *K. pneumoniae* species complex (KpSC), which includes seven species and subspecies that are difficult to distinguish by MALDI-TOF or biochemical tests^30–32^. However, the implications of this population structure to the role of *K. pneumoniae* as an opportunistic pathogen in the hospital setting are unclear, since most WGS studies focus on either MDR (CP or ESBL) HAI, or on hypervirulent community-acquired infections, each of which are associated with a subset of clonal lineages (a few dozen common CP lineages, and fewer than a dozen hypervirulent ones^18,33–35^). WGS has the advantage that it can also be used to identify nosocomial transmission clusters, although this has mainly been applied to investigation of CP or ESBL HAIs^36–39^, or restricted to blood isolates^18,40,41^, and so the overall contribution of nosocomial transmission to total burden of *K. pneumoniae* HAI has not previously been well characterised.

## Methods

### Ethics approval and consent to participate

Ethical approval for this project was granted by the Alfred Hospital Ethics Committee in Melbourne, Australia (Project numbers #550/12 and #526/13). A consent waiver was granted for the inclusion of limited patient data related to clinical isolates, extracted from hospital and laboratory records by hospital staff who normally have access to the data, and shared in deidentified form with research staff for analysis in this project.

### Setting and sample collection

The *Klebsiella* Acquisition Surveillance Project at Alfred Health (KASPAH) was conducted over a one-year period from April 1, 2013 to March 31, 2014 in Melbourne, Australia. All clinical isolates identified as *K. pneumoniae* by the Alfred Health microbiological diagnostic laboratory as part of routine care were included in the study, if they were reported by the laboratory as associated with infection (see details below). Four hospitals in the Alfred Health Network are served by this laboratory. All patients from whose specimens the isolates were cultured were recruited into the study (consent was not required). Relevant clinical data was extracted from the laboratory and hospital records at the time of recruitment (date of specimen collection, specimen type and referring hospital, patient age and gender). Clinical review of hospital records was undertaken retrospectively for all participants, 4 years post-recruitment, in order to classify each isolate as community-acquired (CA), healthcare-associated (HA), or nosocomial in origin (details below).

### Clinical isolates

Clinical isolates were included in the study when the treating physician referred a specimen to the diagnostic service of the microbiology laboratory for analysis based on clinical suspicion of infection, and *K. pneumoniae* was then identified and reported as a pathogen according to the in-house standard operating procedures as previously described^31^. Species identification was performed using matrix-assisted laser desorption ionization-time of flight (MALDI-TOF) (Vitek MS^®^, bioMerieux Marcy l’Etoile, France). All *K. pneumoniae* identified from sterile sites (blood cultures, cerebrospinal fluid, deep tissue biopsies, pleural fluid) and from cultured prosthetic material (e.g., central venous catheters) were reported as pathogens, as long as other enteric or skin flora was not detected. For other specimen types, a *K. pneumoniae* infection was deemed present if sufficient concentrations of neutrophils were seen on microscopy or Gram stain, and *K. pneumoniae* was found to be the sole organism present or the predominant organism if the sample was also expected to contain normal flora. *K. pneumoniae* would be reported as an infection in the absence of neutrophils if the patient was neutropenic. The vast majority of isolates resulted from wound swabs, sputum samples or urine samples. Where *K. pneumoniae* was identified in urine samples or wound swabs along with other enteric bacteria (e.g., *E. coli*), the laboratory reported this as mixed enteric flora; *K. pneumoniae* isolated from such specimens were excluded from the study. Wound swabs were collected when signs of infection were present (i.e., purulent discharge) and reported as *K. pneumoniae* only when other enteric or skin bacteria were not also identified. *K. pneumoniae* was reported as clinically significant when it was the predominant isolate from a well collected sputum specimen. When samples were clearly from the oral cavity (indicated by the presence of saliva (macroscopically), squamous epithelial cells (microscopically) and mixed oral flora on culture), then a small amount of *K. pneumoniae* was not reported as pathogenic but rather as ‘mixed oral/enteric flora’ and such isolates are not included in this study.

Three distinct infection acquisition statuses were defined: community acquired infections, nosocomial infections, and healthcare-associated. Community-acquired (CA) infections were defined by isolation of *K. pneumoniae* from an outpatient or on day 0, 1 or 2 of current admission as an inpatient, and with no recorded prior contact with the Alfred Health Network (either as an inpatient or outpatient) in the previous 12 months. Nosocomial infections were defined by isolation of *K. pneumoniae* on day 3 or later of the current inpatient admission, or with recent inpatient admission (in the last month). *K. pneumoniae* infections in patients not meeting the criteria for nosocomial infection but having some recorded contact with Alfred Health in the last 12 months (including as an inpatient 1-11 months prior to the current infection, or with one or more prior outpatient visits up to 12 months prior to the current infection), were considered as healthcare-associated (HA).

### Antimicrobial susceptibility testing

All clinical isolates were subjected to antimicrobial susceptibility testing in the clinical microbiological diagnostics laboratory upon isolation (i.e., in 2013-2014), using the Vitek2 GN card and interpreted using 2020 EUCAST breakpoints. Antimicrobials tested were: ampicillin (to which *K. pneumoniae* are intrinsically resistant via chromosomally encoded beta-lactamases), amoxicillin-clavulanate, ticarcillin-clavulanate, tazobactam-piperacillin, cefazolin, ceftazidime, ceftriaxone, cefepime, norfloxacin, ciprofloxacin, amikacin, gentamicin, tobramycin, trimethoprim and trimethoprim/sulfamethoxazole. If the susceptibility pattern suggested an ESBL enzyme was present, this was confirmed using the method of Jarlier^42^. Isolates were classified as MDR based on acquired resistance to three or more classes of antimicrobials (i.e. not counting ampicillin resistance which is intrinsic) as previously defined^43^. Selected stored isolates were re-tested in 2021 in order to investigate issues identified upon sequencing: INF034, INF155, INF167, INF255, INF018 (whose genomes were ESBL-negative but ceftriaxone resistant) were re-tested via Vitek2 GN cards; INF307, INF048, INF136 had novel *fosA* genes in the chromosome and were subjected to agar dilution in triplicate to assess MIC to fosfomycin.

### DNA extraction and whole genome sequencing (WGS)

All isolates were subjected to DNA extraction for Illumina sequencing in 2015, using a phenol:chloroform method via phase lock gel tubes (5PRIME) as previously described^31^. Barcoded Illumina DNA libraries were prepared using Nextera XT or TruSeq protocols and sequenced on the HiSeq 2500 platform, generating paired-end reads of 125 bp each. Eighty-seven isolates (26%, see **Table S1**) were later subjected to fresh DNA extraction using a protocol based on GenFind (Beckman Coulter) reagents (doi: 10.17504/protocols.io.p5mdq46), and multiplex long-read sequencing with an Oxford Nanopore Technologies (ONT) MinION device (as described previously^44^), to facilitate assembly, pan-genome and plasmid analyses.

### Species analysis and quality control of WGS data

A total of 362 infection isolates from 318 patients were included in the study. Two of these isolates failed the Illumina library preparation step prior to sequencing. Three of the sequenced read sets were excluded from further analysis because preliminary analysis showed that the sequences were dominated by non*-K. pneumoniae* DNA (two were predominantly *Klebsiella oxytoca* and one was predominantly *Acinetobacter baumannii*). This could be due to either mixed culture with *K. pneumoniae* and another bacterium, or contamination following the initial identification of *K. pneumoniae*. Since the original identification was recorded as *K. pneumoniae*, and the presence of *K. pneumoniae* DNA was confirmed by sequencing, we include these three specimens in the reporting of *K. pneumoniae* clinical isolates; but excluded them from further genomic analysis. A further 29 DNA sequences were excluded from genomic analysis due to either (i) failing quality control thresholds (mean read depth <25×, coverage of reference sequence <85%), or (ii) suspicion of mixed *Klebsiella* strains (ratio of heterozygous:homozygous core gene variant sites ≥2%). The remaining 328 WGS-confirmed *K. pneumoniae* isolates (from 289 patients) underwent detailed genomic analyses. A flow chart of isolate and genome processing is given in **Figure S1**, details of isolates and WGS data accessions are given in **Table S1**.

### Genome assembly, annotation and pan-genome analysis

Illumina short reads were trimmed using Trim Galore v0.5.0 (github.com/FelixKrueger/TrimGalore) (default settings) and assembled *de novo* using SPAdes optimised with Unicycler v0.4.7^45^. Where ONT reads were available, these were processed as described previously^44^ and combined with Illumina reads to generate hybrid assemblies using Unicycler v0.4.7. Contigs were annotated with Prokka v1.13.3^46^. Pan-genome analysis was performed using panaroo v1.1.2^47^ with default settings (sequence identity threshold 95%, protein family sequence identity threshold 70%, length difference threshold 95%).

### Single nucleotide variant analysis and multi-locus sequence typing

Single nucleotide variants (SNVs) were identified by mapping Illumina reads against the ST23 *K. pneumoniae* strain NTUH-K2044 reference genome (NC_012731.1), using the mapping pipeline RedDog v1b.10.3 (github.com/katholt/reddog). RedDog uses Bowtie2 v2.2.5^48^ to map reads and SamTools v1.2^49,50^ to call SNVs with Phred quality score ≥30, as described previously^31^. Multi-locus sequence typing (MLST) was performed, and sequence type (ST) assigned based on the 7-locus scheme^51^, by analysing assemblies with Kleborate v2.0.0^52^. Novel STs were submitted to the *K. pneumoniae* BIGSdb-Pasteur database^23^ for allele assignments. To identify other geographic continents from which STs identified in this study have previously been reported, we used MLST data reported for 13,156 whole genome sequences publicly available in RefSeq in July 2020 (available as Supplementary Data 2 in Lam et al, 2021^52^).

### Phylogenetic analysis

Core genes were defined as those that were annotated in the reference genome and present (coverage ≥95% and mean read depth ≥5×) in all of the sequenced isolates based on the mapping analysis. A maximum likelihood phylogenetic tree was inferred from an alignment of all homozygous SNVs (n=690,727 SNVs) identified within 3,135 core genes in the 328 genomes, using FastTree v2.1.8^53^. The tree file is available via MicroReact (microreact.org/project/gVB3ki6iA62RREC1stLLHo-kaspah-clinical-isolates). Phylogenetic clusters with a maximum pairwise patristic distance of 0.01 (∼6,900 core SNVs, 0.13% nucleotide divergence) were extracted from the trees using R to define lineages.

### Surface antigen biosynthesis and acquired virulence loci

Capsule locus (KL) types and lipopolysaccharide O antigen (O) types were identified from the resulting assemblies using Kaptive v2.0^54,55^; KL and O types with a match confidence of ‘good’ or better (as described at github.com/katholt/Kaptive) were reported; genomes with a match confidence of ‘low’ or ‘none’ were investigated through manual exploration of the assembly graph in Bandage v0.8.1^56^. Putative novel loci were extracted and annotated with Prokka^46^ followed by manual curation. Loci that could not be resolved via manual inspection were marked as “unknown” (i.e., if the assembly graph was fragmented in the region of the K/O locus, or because there was not a single unambiguous path through the locus). Kleborate v2.0.0^52^ was used to screen each genome assembly for key acquired virulence factors that are significantly associated with invasive infections in humans^22^: yersiniabactin^24^ (*ybt*), salmochelin^35^ (*iro*), colibactin^24^ (*clb*), aerobactin^35^ (*iuc*), and regulators of the mucoid phenotype (*rmp* locus, *rmpA2*).

### Plasmid analyses

Plasmid content was assessed using multiple methods. Replicon markers were identified by screening assemblies against the PlasmidFinder database^57^ using BLASTn (80% identity and 80% coverage thresholds). Mob types were assigned using iterative PSI BLAST as described previously^58^. Contigs were assigned as being of plasmid or chromosomal origin using Kraken as previously described^59^ (all other contigs were marked as ‘unknown’).

### Genetic determinants of antimicrobial resistance (AMR)

Kleborate v2.0.0 was used to screen each genome assembly for acquired resistance genes and known chromosomal mutations associated with resistance to fluoroquinolones, colistin and carbapenems^52^. The detected AMR determinants were used to predict resistance to ceftriaxone (based on presence of acquired ESBL genes), meropenem (acquired carbapenemases and *ompK35/36* alleles), ciprofloxacin (known *gyrA* and *parC* mutations and acquired *qnr* genes, but not *aac(6’)-Ib-cr* as there is no evidence this gene can raise the MIC above the breakpoint for clinical resistance in the absence of other determinants^60^), gentamicin (acquired resistance genes defined in CARD v3.0.8^61^), trimethoprim (acquired *dfr* genes), and trimethoprim/sulfamethoxazole (acquired *dfr* genes plus *sul* genes). A major error was defined as a phenotypically susceptible isolate that carried one or more determinants of resistance for the specified drug; a very major error was defined as a phenotypically resistant isolate in which no known resistance determinant for that drug was identified in the genome.

### Detection of species hybrids

The genome collection was screened for hybrids by using BLASTn to align contigs against a set of reference assemblies for each *Klebsiella* species. The BLAST alignments were then used to assign per-species sequence identity to each position in the contig. Each assembly’s overall species composition was then quantified, based on assignment of genomic regions to the closest matching species, and hybrids identified as those with ≥3% of the genome assigned to a second KpSC species. This analysis was implemented in a Python script, available at http://github.com/rrwick/Klebsiella-assembly-species.

### Transmission analysis

Pairwise core gene SNV counts were extracted from the SNV alignment described above, and used to infer transmission networks comprising nodes (one representative isolate per infection episode) connected by edges where the pairwise distance was ≤25 SNVs (based on our previous investigation of within-patient vs between-patient SNV distances^31^, and other recent studies of CP *K. pneumoniae* transmission^38,39^) and the temporal distance was ≤45 days (twice the median of time-to-infection estimated for colonized patients^14^). The *network* function in the R package *network* (v1.17.1) was used to construct the transmission network, and to extract clusters of connected nodes (isolates). Putative transmission clusters were manually reviewed for plausible epidemiological links; one was removed because onset of the second case occurred on day 1 of admission and no previous admissions with Alfred Health were recorded; another was removed because it comprised specimens taken from one outpatient and one inpatient collected on the same day.

### Statistical analysis

All statistical analyses were conducted using R (v3.6.3). Specific tests used are given together with each result in the text, corresponding R functions are: *wilcox*.*test* for Wilcoxon rank-sum test; *prop*.*test* for test of differences in proportions; *fisher*.*test* for Fisher’s exact test; *chisq*.*test* for Chi-square test; *bartels*.*rank*.*test* with left-sided test for trend, to test for temporal trends in monthly isolate counts; *glm* with ‘family=binomial(link=‘logit’)’ for logistic regression. Figures were plotted in R using *ggplot2* v3.3.5, *ggnetwork*^62^ v0.5.10 and *ggtree*^63^ v2.4.2.

## Results

### Infection burden

A total of 362 clinical isolates of KpSC were identified at the microbiological diagnostic laboratory during the 1-year study period, collected from 318 patients (**Figure 1, Table 1**). The patients were 55% female and ranged in age from 20 to 97 years old, with median age 70 years. The median age for females was significantly higher than for males (75 vs 67, p=0.001 using Wilcoxon rank-sum test, see **Figure 1d**). The majority of patients had UTI (66%), 15% had pneumonia and 10% wound/tissue infections (**Figure 1a, Table 1**). Ten percent of patients had disseminated infections (bloodstream and/or cerebral spinal fluid (CSF) isolates); most had no other isolate and the primary site of infection was not known (**Table 1**). UTIs were more common in females whilst pneumonia was more common in males (see **Figure 1d, Table 1**). No statistically significant differences were observed in gender or age distribution for other infection types (**Table 1**). KpSC clinical isolates originated from specimens taken in 49 clinical units/wards; those contributing the greatest number of isolates were the emergency department (n=72, majority causing UTIs (n=54) plus 12 disseminated infections), ICU (n=41, majority causing pneumonia (n=25) plus four disseminated infections), and haematology ward (n=15, majority causing pneumonia (n=5) or disseminated infections (n=7)). Sixteen isolates were collected in outpatient clinics (n=15 UTI, n=1 pneumonia).

**Figure 1.**
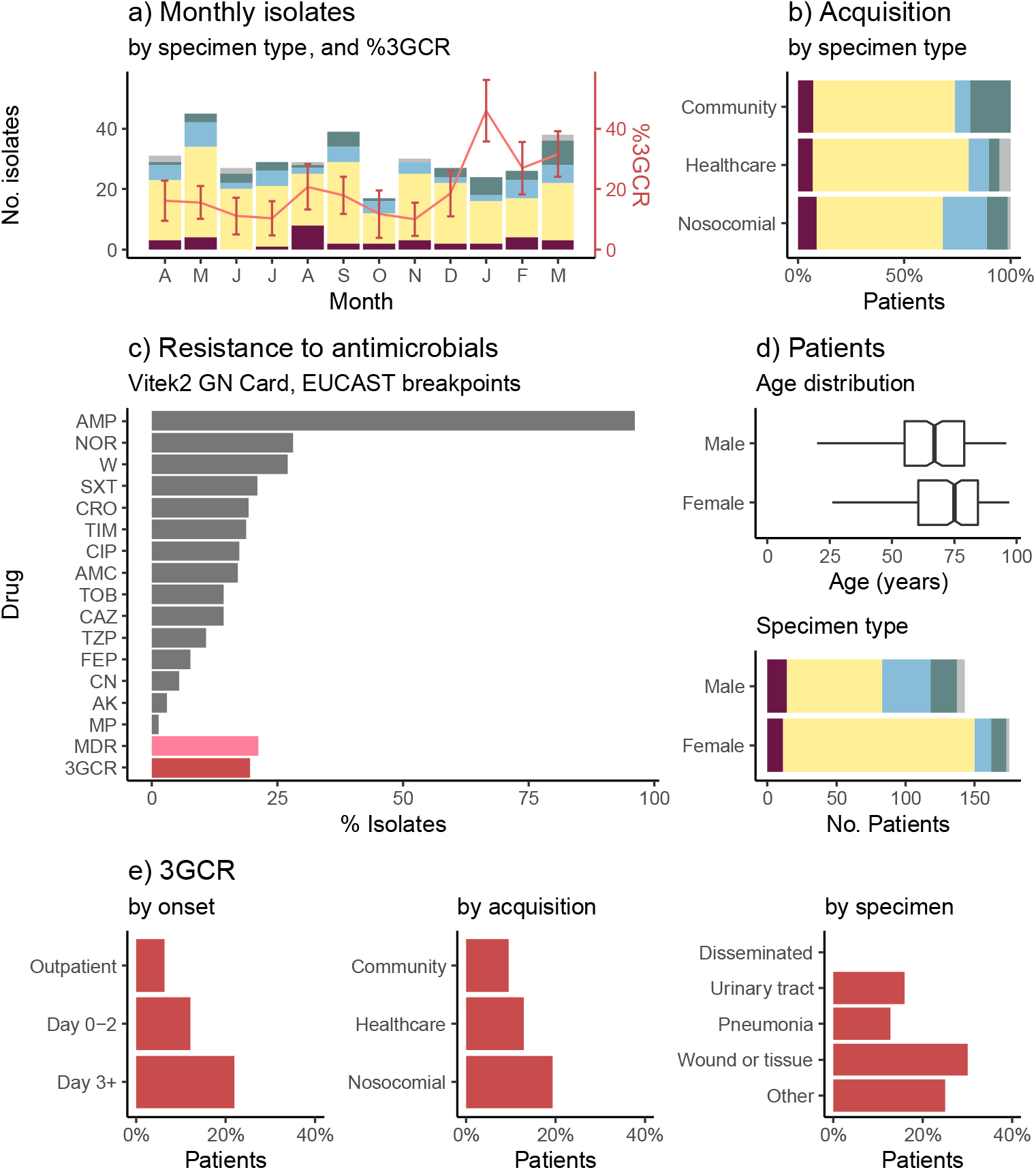
Characteristics of clinical isolates identified as *K. pneumoniae* in the hospital microbiological diagnostic laboratory. **(a)** Monthly isolate counts (total 362 isolates, from 318 patients), coloured by specimen type (maroon=disseminated, yellow=urine, blue=respiratory, green=wound, grey=other). Red line shows frequency of 3^rd^ generation cephalosporin resistant (3GCR) isolates per month, according to the right-hand y-axis (error bars indicate standard error). **(b)** Specimen types (coloured as per panel a) according to inferred mode of acquisition of infection (inferred from hospital contact in past 30 days (nosocomial) or past year (healthcare associated), see **Methods**). **(c)** Bars show proportion of isolates resistant to each drug (AK, amikacin; AMC, amoxicillin–clavulanic acid; AMP, ampicillin; CAZ, ceftazidime; CIP, ciprofloxacin; CRO, ceftriaxone; FEP, cefepime; CN, gentamicin; MP, meropenem; NOR, norfloxacin; SXT, trimethoprim-sulfamethoxazole; TIM, ticarcillin–clavulanic acid; W, trimethoprim; TOB, tobramycin; TZP, tazobactam-piperacillin). MDR=resistant to ≥3 drug classes other than ampicillin; 3GCR=resistant to CRO and/or CAZ. **(d)** Characteristics of patients (n=318) from whom *K. pneumoniae* was isolated. Age distribution: points indicate individual patients (those with ≥1 3GCR isolate are coloured red), boxplots summarise median (central line), interquartile range (boxes). Specimen type: stacked bars are coloured as per panel (a) to indicate the first specimen type, for each patient, from which *K. pneumoniae* was isolated. **(e)** 3GCR rates (amongst first isolate per patient, n=318 patients), stratified by (i) day of onset of infection, relative to day of hospital admission=0; (ii) mode of acquisition, inferred from hospital contact in past 30 days (nosocomial) or past year (healthcare associated) (see **Methods**); (iii) specimen type (note one patient had 3GCR disseminated infection, after an initial susceptible UTI).

**Table 1.** Characteristics of patients with KpSC clinical isolates. UTI=urinary tract infection; Acquisition refers to likely mode of acquisition of infection, inferred from hospital contact in past 30 days (nosocomial) or past year (healthcare associated), see **Methods**. **p<1.4×10^-8^, *p=1×10^-5^ for association between sample type and female sex (using Fisher’s exact test).

|  | **Total**<br>**N (% of total)** | **Female**<br>**N (% of row)** |
| --- | --- | --- |
| **All patients** | **318 (100%)** | **175 (55%)** |
| **Infection type/s** |  |  |
| UTI only | 202 (64%) | 135 (67%) ** |
| + disseminated | 6 (2%) | 4 (67%) |
| + wound/tissue | 2 (0.6%) | 2 (100%) |
| Respiratory only | 46 (15%) | 12 (26%) * |
| + disseminated | 1 (0.3%) | 0 |
| Disseminated only | 24 (8%) | 10 (42%) |
| Wound/Tissue only | 29 (9%) | 10 (34%) |
| Other | 8 (3%) | 2 (25%) |
| **Onset (first clinical isolate, relative to day of first hospital admission)** |  |  |
| Day 0-2 | 175 (55%) | 105 (60%) |
| Day 3+ | 127 (40%) | 59 (47%) |
| Outpatient | 16 (5%) | 11 (69%) |
| **Acquisition (first clinical isolate per patient)** |  |  |
| Nosocomial | 157 (49%) | 76 (48%) |
| Healthcare | 118 (37%) | 72 (61%) |
| Community | 42 (13%) | 26 (62%) |
| **Hospital** |  |  |
| Hospital A | 219 (69%) | 109 (50%) |
| Hospital C | 40 (13%) | 25 (63%) |
| Hospital D | 34 (11%) | 25 (74%) |
| Hospital B | 9 (3%) | 5 (56%) |
| Outpatients | 16 (5%) | 11 (69%) |

Only 40% of patients had their first clinical KpSC isolate collected >2 days into their current hospital admission (i.e. meeting the standard definition of nosocomial onset) (**Table 1**). Taking into account prior contact with the hospital network in the last 1-12 months to ascertain likely mode of acquisition (nosocomial: onset on day >2 and/or prior hospital admission within 30 days; HA: hospital or outpatient contact in last 12 months and onset on day ≤2; CA: no such contact and onset on day ≤2; see **Methods** for details), we estimate that 49% of KpSC infections were nosocomial and a further 37% were HA. Just 13% of infections could be considered CA, of which 67% were UTIs and 19% wound infections (**Figure 1b**). Forty percent of CA infections were admitted via the emergency department (n=12 UTI, 3 disseminated and 2 wound infections) and 12% via ICU (n=4 wound infections and 1 pneumonia). Pneumonia was significantly more common amongst nosocomial acquired infections (21%; vs 9.5% of HA and 7.1% of CA, p=0.005 for test of difference in proportions), whilst wound infections were significantly more common amongst CA infections (19%; vs 5% of HA and 10% of nosocomial, p=0.045 for test of difference in proportions) (see **Figure 1b**).

The frequencies of AMR phenotypes per KpSC isolate are shown in **Figure 1c**. Most isolates (63%) were susceptible to all drugs tested except ampicillin. The remaining 37% had acquired resistance to ≥1 drug tested, 21.3% were MDR (acquired resistance to ≥3 drug classes), and 19.6% were 3^rd^ generation cephalosporin resistant (3GCR, of which 96% were MDR). At the patient level, four patients (1.4%) had ≥1 carbapenem (meropenem) resistant isolate and 46 (16%) had ≥1 3GCR isolate. 3GCR KpSC infections (using first clinical isolate per patient) were significantly associated with nosocomial infection, whether defined as onset >2 days after admission (OR 1.12 and p=0.01, vs day 0-2 and adjusting for patient age, sex, specimen type using logistic regression), or accounting for other recent admissions (OR 1.13 and p=0.05, vs CA and adjusting as above). Whilst no temporal or seasonal trends in monthly isolation rates were detected, either for individual infection types or in aggregate (p>0.1 using Bartel’s rank test), the 3GCR frequency showed an increasing trend (p=0.036 using Bartel’s rank test for trend, see **Figure 1a**), rising from mean 15% (range 10-21% per month) in the first nine months of the study to mean 34% (27-46% per month) in the final three months (p=0.0002, using test of difference in proportions of 3GCR between the two periods).

### Genomic diversity

All clinical isolates identified as KpSC were subjected to WGS, yielding 328 pure isolate genomes from 289 patients for detailed comparative genomic analysis (**Methods, Figure S1**). WGS confirmed the majority of pure-culture isolates were *K. pneumoniae* (82.3%); the rest were *K. variicola* subsp. *variicola* (13.7%), *K. quasipneumoniae* subsp. *similipneumoniae* (3.7%) and *K. quasipneumoniae* subsp. *quasipneumoniae* (n=1) (**Table S2, Figure 2A**). There were no significant associations between species and infection type, onset or acquisition; however, *K. pneumoniae* infections were more likely to display acquired resistance phenotypes compared to the other taxa combined (32.9% vs 14.5%; p=0.01 using test for difference in proportions) (see **Table S2**).

**Figure 2.**
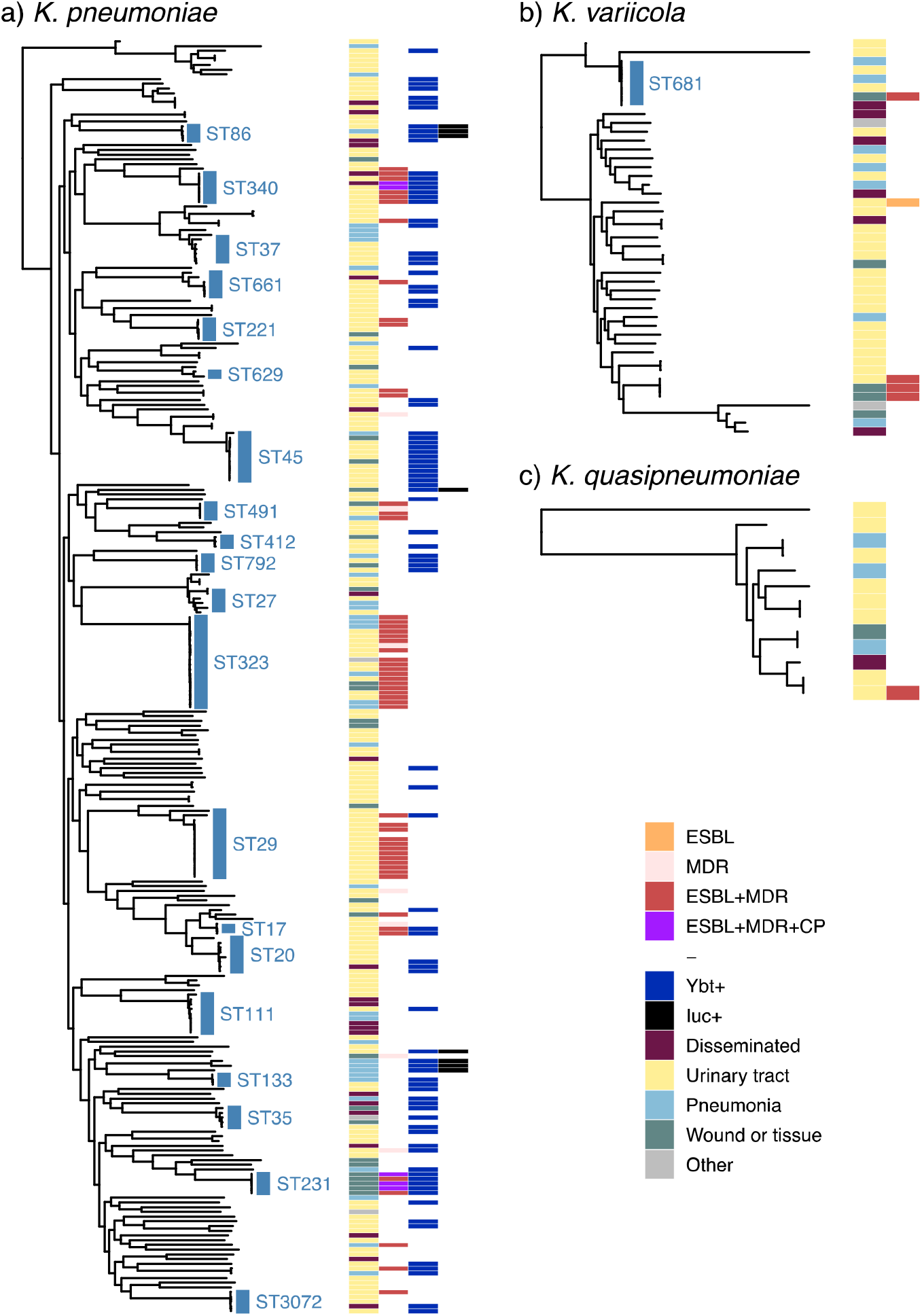
Core genome phylogenies for *K. pneumoniae* species complex isolates. *(a) K. pneumoniae*, (b) *K. variicola*, (c) *K. quasipneumoniae*. Trees shown are subtrees extracted from a maximum likelihood phylogeny inferred from a complex-wide alignment of core gene SNVs for all 328 sequenced clinical isolates (i.e. each species tree is rooted using the others as outgroups), available at <u>microreact.org/project/gVB3ki6iA62RREC1stLLHo-kaspah-clinical-isolates</u>. The 21 ‘common’ lineages, each identified in ≥3 patients, are labelled in blue with their sequence type (ST). Columns indicate (1) infection type (disseminated, urinary tract, respiratory, wound/tissue, other); (2) antimicrobial resistance (ESBL, MDR, ESBL+MDR, ESBL+MDR+CP); (3) yersiniabactin (Ybt+, identified in *K. pneumoniae* only); (4) aerobactin (Iuc+, identified in *K. pneumoniae* only); coloured as per inset legend. ESBL=extended spectrum beta-lactamase, MDR=resistant to ≥3 drug classes other than ampicillin, CP=carbapenemase producing.

We assessed genomic diversity of the clinical isolates in terms of phylogenetic lineages, gene content, plasmid content, AMR and acquired virulence determinants, and surface antigen synthesis loci. We inferred a maximum likelihood core-genome phylogeny (**Figure 2**) and used this to cluster the 328 genomes into 182 lineages (138 *K. pneumoniae*, 35 *K. variicola*, 9 *K. quasipneumoniae*) representing distinct strain types that have been separated from other lineages over many years of evolution (see **Methods**). These correlated very closely with STs defined by 7-locus MLST (see **Figure 2, Figure 3a, Table S1**). Twenty-six patients contributed more than one sequenced isolate. In most cases (n=21, 81%), isolates from the same patient matched at the lineage level, consistent with a single infecting strain and we classified this as a single infection episode. Of the remaining cases, three patients had one lineage identified in urine followed by a disseminated infection with a second lineage 2-19 days after; one patient had different MDR lineages (ST347 and ST491) detected in wound swab specimens collected from the same site three days apart; and one patient had one lineage (sensitive ST520) isolated from sputum and a second lineage detected 32 days later in both blood and sputum (norfloxacin-resistant ST111). We therefore classified these five patients as each having two distinct infection episodes, bringing the total number of genomically-defined infection episodes to 294. The cumulative counts of infection episodes, lineages and STs during the study are plotted in **Figure 3a**. All curves were nearly linear (linear regression, adjusted R^2^≥ 0.97), and the slopes of the lineage/ST linear regression curves were 69% that of total infection episodes (67% considering *K. pneumoniae* only, dashed lines in **Figure 3**), indicating extensive diversity of strains underlying the total infection burden.

**Figure 3.**
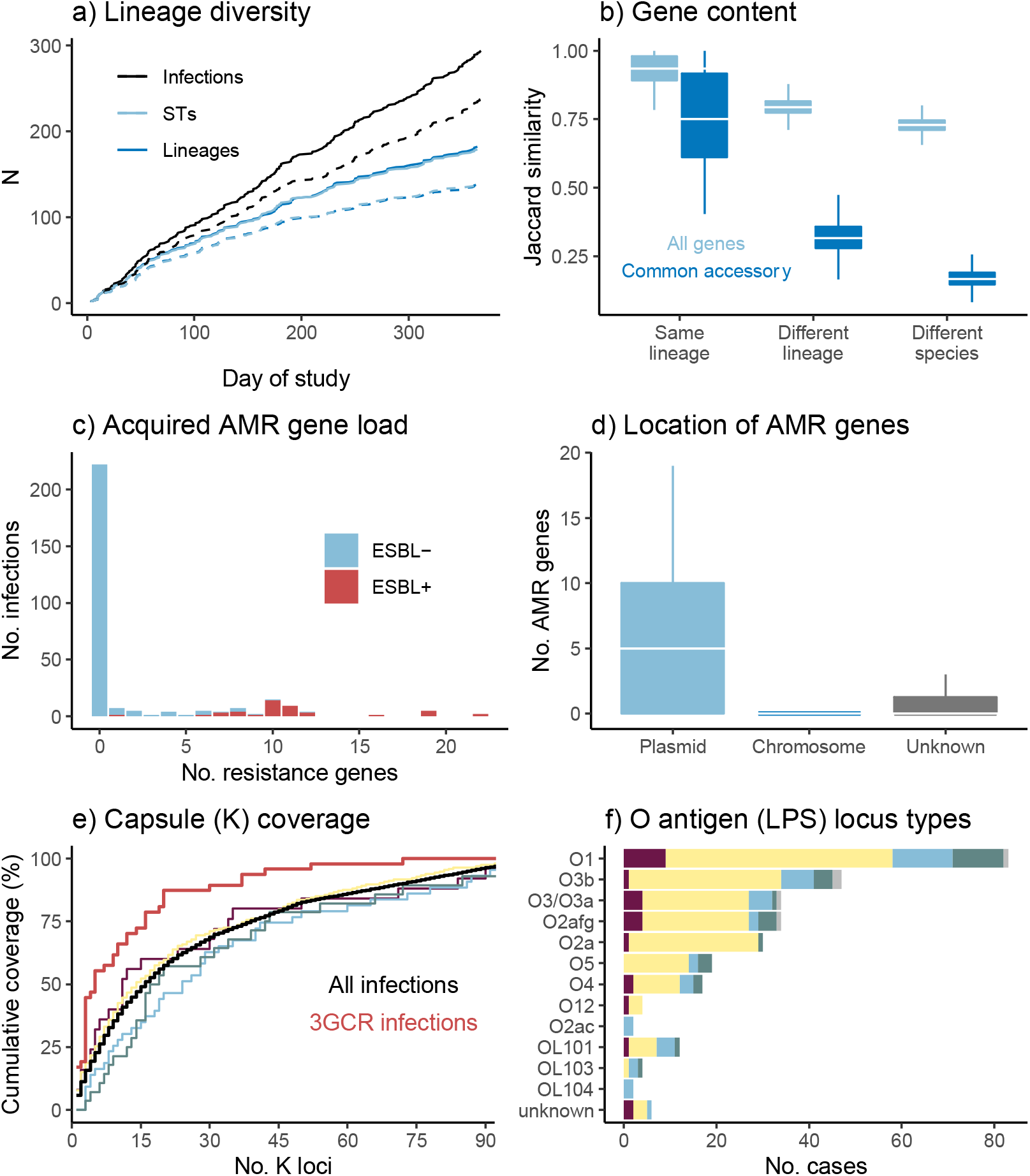
Genomic diversity of clinical isolates identified as *K. pneumoniae* in the hospital microbiological diagnostic laboratory (one per infection episode). **(a)** Accumulation of unique infection episodes (total 294 in 289 patients), lineages or sequence types (STs) over the study period. Solid lines=all *K. pneumoniae* species complex (KpSC); dashed lines=*K. pneumoniae* only (total n=238). **(b)** Distribution of pairwise gene content Jaccard similarity between genomes, calculated using either all genes (including n=3,095 core genes present in all strains), or the n=4,001 common accessory genes (each present in 5-95% of genomes sequenced). **(c)** Distribution of the number of acquired antimicrobial resistance (AMR) genes per genome, stratified by detection of an extended spectrum beta-lactamase (ESBL) gene. **(d)** Number of AMR genes per genome that were attributed to plasmid-or chromosome-derived contigs using Kraken. **(e)** Theoretical coverage of infections (y-axis) by vaccines targeting capsular antigens encoded by increasing numbers of K loci (x-axis, K loci ordered from most to least common in this data set as shown in **Table S4**). Black line shows cumulative coverage of all unique infections (n=294); red line shows unique 3GCR infections (n=47); other lines show coverage of different infection types (maroon=disseminated, yellow=urine, blue=respiratory, green=wound, grey=other). **(f)** Frequency of O antigen (LPS) synthesis loci, stratified by specimen type (coloured as per panel e).

#### Gene content

The mean number of genes per genome was 5,031 (IQR 4,936-5,161), including 3,095 core genes that were present in all genomes. The pangenome comprised 23,075 genes in total, of which 4,067 were very common (present in ≥95%), 4,001 were common (present in 5–95%), and 15,007 were rare (present in <5%) or very rare (9,845 in <1%) (**Figure S2**). Gene content was largely conserved within lineages (median pairwise Jaccard similarity 0.93 for all genes including core genes, and 0.75 across common genes) but was quite distinct between lineages of the same species (median values 0.79 and 0.32 for all genes and common accessory genes, respectively) (**Figure 3b**). We used multiple methods to assess plasmid load and diversity. *Mob* markers were detected in genomes from 89% of infection episodes (median n=2, IQR 1-3), and *rep* markers were detected in 84% (median n=5, IQR 2-9). A total of 55 uniquely distributed *rep* markers were identified, including 25 that were present in ≥5% of infection episodes and 18 in ≥10% (**Figure S3**). The number of unique *mob* and *rep* markers were significantly correlated across genomes (Pearson correlation coefficient=0.614, p<1×10^-15^), and both were significantly independently associated with total DNA in contigs predicted to be plasmid-derived (see **Figure S3**). In total, 252 infection episodes (86%) had predicted plasmid load ≥5 kbp (89% with >0 bp, 85% with ≥10 kbp). Amongst these presumptive plasmid-positive genomes, median plasmid load was 233 kbp (range 7.3-704 kbp, IQR 171-306 kbp).

#### AMR determinants

Screening for known AMR determinants identified 60 acquired AMR genes in genomes from 73 infection episodes (24.7%), including 44 (15%) carrying ESBLs and 5 (1.7%) with carbapenemases. Seven distinct fluoroquinolone resistance-associated *gyrA*/*parC* mutations were detected in genomes from 18 infections (6%; including n=15 that also had acquired AMR genes). The number of acquired AMR genes per isolate was bimodal; amongst those with any acquired AMR genes, the median was 10 genes and the interquartile range (IQR) 6-11 genes (range 1-22) (see **Figure 3c**). ESBL genes were concentrated in 15 lineages (8%; 13 *K. pneumoniae*, 2 *K. variicola*; see **Figure 2**). Six lineages accounted for 82% of ESBL infections (corresponding to ST323, ST29, ST491, ST340, ST231, ST17). Most (98%) ESBL-positive genomes carried other acquired AMR genes (median 10, IQR 9-11; see **Figure 3c**), consistent with the high rate of the MDR phenotype amongst 3GCR isolates (96%). Carbapenemases were identified in *K. pneumoniae* ST340 (n=2 infections, *bla*_IMP-4_) and ST231 (n=3 infections, *bla*_OXA-48_). The majority of acquired AMR genes were predicted to be plasmid-borne (67.5%), and 8% chromosomally located (the rest were unassignable, **Figure 3d**). Chromosomally integrated AMR genes were identified in genomes from 12 infection episodes and confirmed by long read sequencing (**Table S1**). These included six genomes (three ST231, three ST340) with *bla*_CTX-M-15_ integrated in the chromosome via IS*Ecp1*, and one (ST29) with an entire 243 kbp MDR plasmid fused with the chromosome as previously reported^12^. Three chromosomes (*K. variicola* ST616 and ST1456) carried a novel acquired *fosA* homolog (closest relative being *fosA7*, 9% nucleotide divergence); however only one of these isolates (INF136) had elevated fosfomycin MIC (128 mg/L, vs wildtype range 16-32 mg/L) so it is not clear whether this gene confers resistance.

Concerningly the 16S rRNA methylase gene *rmtB* (which confers high-level resistance to aminoglycosides) was found in isolates from three patients, displaying resistance to amikacin, gentamicin and tobramycin in addition to 3GC, amoxicillin-clavulanic acid, ticarcillin-clavulanic acid, tazobactam-piperacillin, ciprofloxacin, norfloxacin and trimethoprim/sulfamethoxazole. One was resistant to meropenem (MIC ≥16 mg/mL) and the other two had elevated MIC (1 mg/mL, compared to the wildtype <0.25 mg/mL for 96.4% of isolates). These three isolates were all ST231 and harboured three quinolone-resistance mutations (GyrA-83I, GyrA-87Y, ParC-80I) and a *bla*_CTX-M-15_ insertion in the chromosome; an IncC plasmid carrying *rmtB* plus 11 other AMR genes including the ESBLs CTX-M-15 and VEB-1, *ermB* (azithromycin resistance) and *arr-2* (rifampicin resistance); and an IncL plasmid carrying the OXA-48 carbapenemase.

AMR phenotypes for key drugs were quite well predicted by known AMR determinants, however there were instances of unexplained resistance for most drugs (**Table 2**). Of the 47 3GCR infections, 42 carried known ESBL genes (n=33 *bla*_CTX-M-15_ (n=3/33 also carried *bla*_VEB-1_), n=6 *bla*_CTX-M-14_, n=1 *bla*_CTX-M-3_, n=1 *bla*_CTX-M-62_, n=1 *bla*_SHV-12_). The remaining five isolates were MDR and had tested positive for ESBL production in the diagnostic laboratory, but the corresponding genome sequences lacked ESBL genes and four lacked any acquired AMR genes. Re-testing of stocked cultures confirmed that four remained resistant to ceftriaxone (and were MDR), but one (INF255) had regained susceptibility to cephalosporins, fluoroquinolones, aminoglycosides and trimethoprim/sulfamethoxazole. Another (INF018) was an ST323 separated from other ESBL-positive ST323 strains by ∼30 SNVs. We therefore speculate that all five isolates initially had ESBL/MDR plasmids upon first isolation and testing, but these were lost during culture for DNA extraction (this would also account for most unexplained resistance for other drugs except trimethoprim).

**Table 2.**
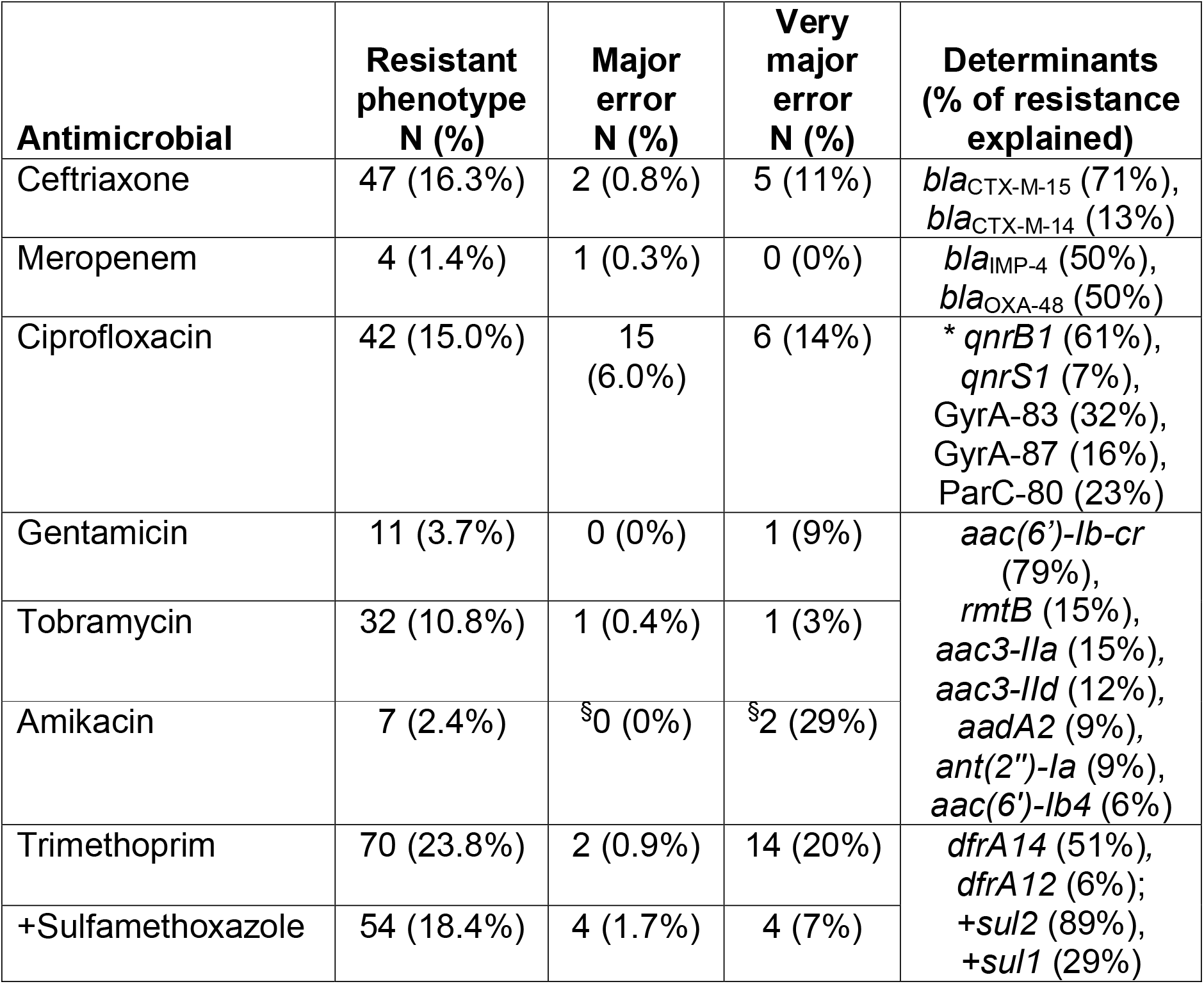
Comparison of AMR phenotype vs genotype. Based on most resistant isolate per infection episode (n=294). Major error = number (%) of susceptible isolates in which a resistance determinant was identified; very major error = number (%) of resistant isolates in which a resistance determinant was not identified (i.e. unexplained resistance). Genetic determinants explaining ≥5% of resistance to the given drug or class are listed. *Note *aac(6’)-Ib-cr* reduces susceptibility to fluoroquinolones (Machuca *et al*, 2016) but does not on its own raise the MIC above the breakpoint for clinical resistance; it was present in 60% of ciprofloxacin resistant strains, but these all carried *qnr* genes and/or *gyrA*+*parC* mutations that could explain resistance; hence *aac(6’)-Ib-cr* is not included in the list of genetic determinants explaining ciprofloxacin resistance. ^§^Errors for amikacin resistance prediction (MIC ≥16 mg/L) shown in the table are based on excluding *aac(6’)-Ib-cr* as a determinant, as we could not find any evidence that this specific allele confers resistance in *Klebsiella* (causative genes detected were *rmtB* and *aac(6’)-Ib4*); including *aac(6’)-Ib-cr* removes the two very major errors but results in 24 major errors due to isolates carrying the gene but which have low MIC for amikacin (MIC 8 (n=2), 4 (n=7) or <2 (n=15)).

#### Acquired virulence determinants

The *ybt* locus encoding the acquired siderophore yersiniabactin was detected in isolates from 33% of *K. pneumoniae* infection episodes associated with 48 lineages (**Figure 2**) but was not detected in other KpSC. Fourteen *ybt* locus types were identified (including the plasmid-borne *ybt* 4 in 14 strains from 11 STs, as previously reported^24^). Presence of *ybt* was not significantly associated with the presence of ESBL or other acquired AMR genes, however the additional acquired virulence factors *iuc, iro, rmp* and *clb* were exclusively found in *ybt*+ isolates, with overall frequencies of 2.7% (*iuc*) or 3.1% (*iro, rmp, clb). Iuc, iro* and *rmp* were mostly concentrated together in five known hypervirulent clonal groups, associated with virulence plasmids (CG86, CG23, CG66, CG420, CG91/subsp. *ozaenae*; **Table S3**). Isolates harbouring the complete *rmp* locus were confirmed to be hypermucoid via the string test (except for a single ST86 isolate). Three *clb* variants were identified in five STs (**Table S3**), located with *ybt* in ICE*Kp10*.

#### Surface antigens

Capsule (K) and LPS (O) antigens have been proposed as targets for vaccination against *K. pneumoniae* infection^64^. K biosynthesis loci were confidently identified for n=284 (96.6%) infection episodes, spanning 91 distinct K locus (KL) types (including 4 novel loci, KL167-170, see **Table S4**). Forty-one K loci (45%) were identified only once each (**Table S4**). Eight KL types were found in ≥3% of infection episodes each (**Figure S4**), together these accounted for 33% of all infection episodes including 57% of 3GCR cases (**Figure 3e)**. These top eight KL types were each associated with a dominant ST (**Figure S4**), suggesting that their comparatively high prevalence was driven by local clonal expansion (and potentially transmission) of specific lineages. However, all except one (KL21) were also found in multiple STs (4-7 STs each) indicating that these K types are also widely distributed across lineages (**Figure S4**). Operons for synthesis of the most common capsular polysaccharide sugar components, mannose (*man*) and rhamnose (*rml*), were present in the K loci of 64% and 29% of unique infection episodes respectively (**Table S4**); these were not significantly associated with infection type (p=0.71 and p=0.19, respectively, using Chi-square test).

The theoretical coverage provided by multi-valent vaccines targeting increasing numbers of KL (ordered by KL frequency in the population) is shown in **Figure 3e**. KL diversity was similar for each type of infection (Simpson’s diversities between 0.93 and 0.97), and theoretical vaccine coverage was also similar (**Figure 3e**). Overall, 16 KL types (each with frequency ≥2%) would need to be targeted to cover 50% of infections (79% of 3GCR), and 31 KL types (each with frequency ≥1%) to cover 70% of infections (89% of 3GCR) (**Figure 3e**). Enhanced coverage of 3GCR infections is attributable to high numbers of the ESBL-producing strains ST323 (KL21) and ST29 (KL30), which were transmitted in the hospital (discussed below).

Twelve distinct O types were predicted amongst 284 infections with typeable O loci (96.6%), with similar diversity observed within each of the four most common infection types (Simpson’s diversities between 0.75 and 0.83) (**Figure 3f**). The most common O types were O1, O2 (including subtypes O2afg, O2a and O2ac) and O3 (including subtypes O3a and O3b), which together accounted for 77% of infection episodes (including 70% of 3GCR); O1 and O2 accounted for 49% (36% of 3GCR). The common 3GCR STs were O3b (ST323) and O1 (ST29). The mannose-containing O types (O3, O5, OL104) accounted for 35% of infections, but were not associated with infection site (p=0.33 using Chi-square test).

#### Species hybrids

As hybridization between KpSC members has been reported previously^22^ we screened our genome collection for evidence of cross-species hybridization (see **Methods**) and identified 12 *K. variicola* clinical isolates whose genomes harboured imports of between ∼100 kbp and ∼1 Mbp of sequence from *K. pneumoniae*. These represent 12 unique infection episodes and 8 lineages, i.e., 27% of *K. variicola* infections and 23% of *K. variicola* lineages. Four further hybrids were identified amongst isolates reported previously from screening swabs at the same hospitals^12,31^

Ten of the hybrids belonged to *K. variicola* ST681 (3 UTI, 2 respiratory, 1 disseminated, 1 unknown, 3 throat swabs). One clinical respiratory isolate and two subsequent screening throat swabs were isolated from a single ICU patient. Genomic comparisons with publicly available ST681 genomes suggest that our ST681 isolates were in fact hybrids of *K. variicola* ST681 and *K. pneumoniae* (see **Methods** and **Figure S5**). One isolate (INF232; from a woman in her 90s with UTI at Hospital D) comprised a ST681 *K. variicola* genome backbone with a 281 kbp recombinant region whose sequence closely matched *K. pneumoniae* (≤0.5% divergence, **Figure S5a**). This isolate harboured KL143 (*man*-positive) and a truncated form of the O3/O3a locus (broken by an insertion of IS903B and likely non-functional). The nine other local ST681 isolates were very closely related to one another (0-7 pairwise SNVs) and shared with INF232 the 281 kbp *K. pneumoniae* import and also a second recombination import of 311 kbp, which spanned the K and O biosynthesis loci and resulted in import of intact KL10 (*man*/mannose-positive) and O1/O2v2 (O2afg) loci. Again, the imported region showed close homology (≤0.5% divergence) with *K. pneumoniae*, in which KL10 was originally described (**Figure S5a**).

The other six hybrids all included recombinant regions spanning the K locus, resulting in import of various capsule loci from *K. pneumoniae* (see **Figure S5B-d**). Five were associated with infections (UTI, respiratory, wound, sepsis) and one was from a rectal screening swab in the ICU study^31^. Four of these hybrids belonged to ST925, each comprising a *K. variicola* ST925 backbone with a different recombinant block spanning the K locus, between 90 and 565 kbp in size, apparently imported from *K. pneumoniae* (≤0.5% divergence; see **Figure S5b**) and encoding distinct K and O types (KL9/O1, KL28/O3, KL102/O1, KL169/OL104). These strains, isolated from four different patients at Hospital A, differed from one another by >12,000 SNVs in the non-recombinant backbone regions, confirming they were not related to one another by recent local transmission. The other two hybrids were novel singleton STs (ST3095 and ST3060), also comprising *K. variicola* with one or two imported regions from *K. pneumoniae* (393 to 1,043 kbp in size, **Figure S5c-d**).

### Genomics-informed understanding of disease burden

Of the total 182 lineages associated with 294 unique infection episodes, 139 (76.4%) were unique to an individual patient. These singleton lineages accounted for nearly half (47.3%) of all the infections, which most likely originate from the patients pre-existing gut microbiome. The remaining infection episodes were associated with 43 lineages that were detected in multiple patients, including 21 ‘common’ lineages (11.5% of all detected) that were each isolated from ≥3 patients and accounted for 37.8% of all infection episodes (labelled in **Figure 2**). These comprised 20 *K. pneumoniae* and the hybrid *K. variicola* ST681 cluster. Isolates belonging to the common lineages were significantly and independently positively associated with ESBL, *ybt, man*-positive K loci, and nosocomial onset; and negatively associated with non-*K. pneumoniae* species and *rml*-positive K loci (**Figure 4a**).

**Figure 4.**
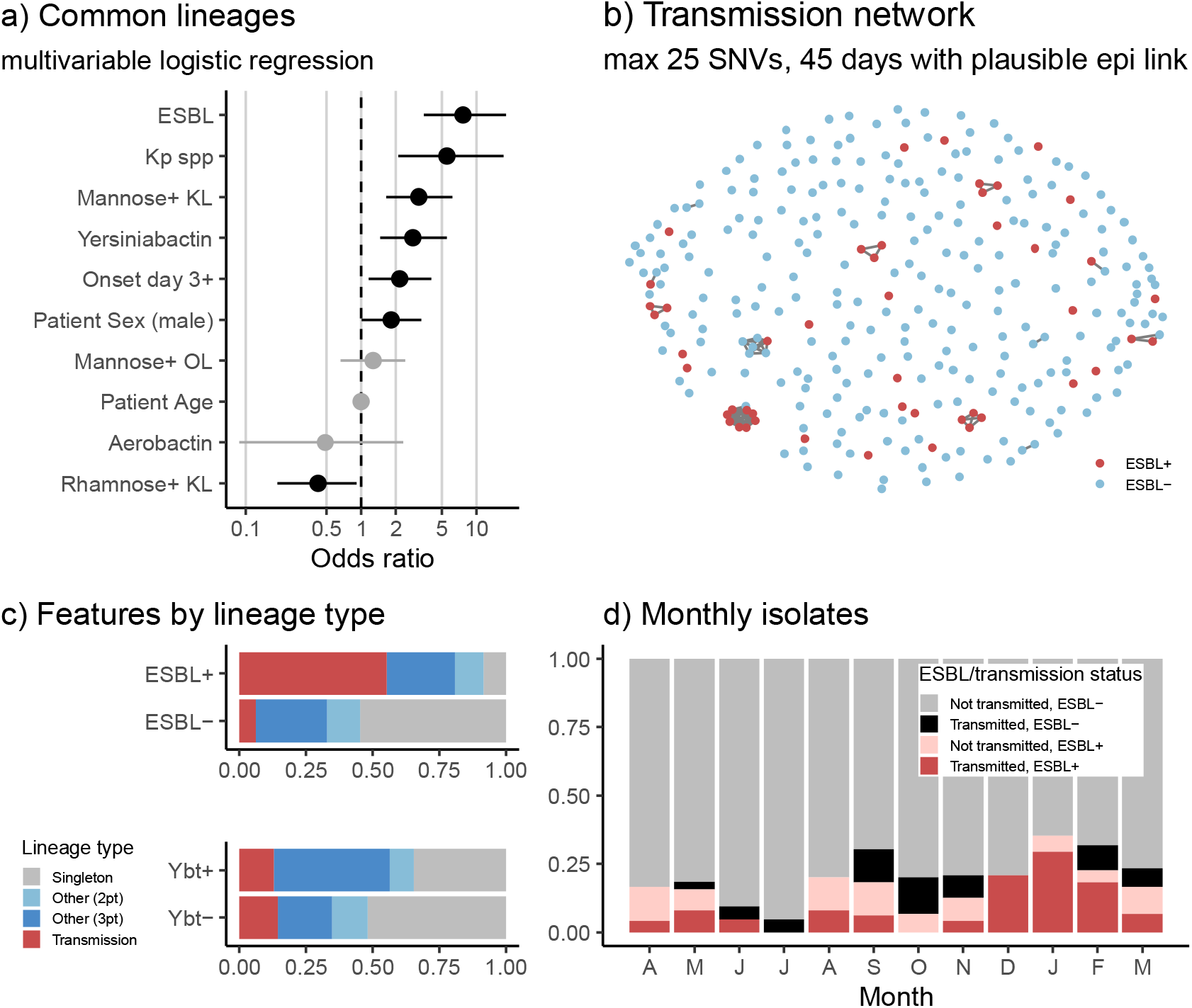
Features of common lineages, and contribution of common and transmitted lineages to infection burden. (a) Genetic and patient characteristics associated with common lineages (identified in ≥3 patients). Circles indicate odds ratios estimated in a single multivariable logistic regression model with all 10 predictors; lines indicate 95% confidence intervals for those odds ratios; significant (p<0.05) associations are coloured black. Binary variables: ESBL (extended spectrum beta-lactamase detected); Kp spp (*K. pneumoniae* species); Mannose+ KL (K locus includes *man* operon); Yersiniabactin (*ybt* detected); Onset day 3+ (isolated from specimen collected on day 3 or later); Patient sex (male); Mannose+ OL (O locus includes *man* operon); Aerobactin (*iuc* detected); Rhamnose+ KL (K locus includes *rml* operon). Continuous variable: Patient age (years). (b) Transmission network showing 12 clusters, defined as infection episodes in different patients identified less than 45 days apart with isolate genomes separated by ≤25 SNVs (mean 0.7 SNVs) and with plausible epidemiology (onset of non-index case occurred during hospital stay, at least 2 days after index case, and patients spent time in same hospital). Details of clusters are given in **Table S5**. (c) Infections of different classes (ESBL+/-, ybt+/-) stratified by lineage type (transmission cluster, as defined in b; other common lineage, identified in ≥3 patients; lineage identified in 2 patients; or singleton lineage identified in 1 patient). (d) Monthly frequencies of ESBL+ and/or transmission-linked isolates. These analyses were completed using one isolate per unique infection episode (n=294); where there were ESBL+ and ESBL-variants of the same strain associated with the same infection episode, the ESBL+ variant was included in order to best reflect the nature of the ESBL infection burden.

#### Burden and risk factors of nosocomial transmission

The frequency of the common lineages could potentially reflect nosocomial transmission, or a higher propensity to cause disease in colonized patients. We defined probable nosocomial transmission clusters as those with ≤25 pairwise SNVs between genomes isolated from the same hospital within 45 days and with plausible epidemiological links (**Figure 4b**, see **Methods**). This identified 12 clusters of 2-9 patients each, involving 11 STs and associated with 41 infection episodes (14%) (**Table S5**). Mean pairwise distance between clustered isolates was 0.7 SNVs (median 0, IQR 0-0, range 0-22). As expected, these infections were significantly associated with onset several days into the hospital stay (median onset day 4 for infections in transmission clusters, vs day 1 for other infections, p=0.007 using Wilcoxon test). Notably one of these clusters involved the ST681 hybrid strain, which infected six ICU patients over a 2.5-month period. Patient age was independently associated with transmission (OR 0.98 [95% CI, 0.96-0.996], p=0.02 in multivariable logistic regression model); but patient sex and bacterial virulence factors were not (**Table S6**). Two-thirds of the transmission clusters involved ESBL+ strains (**Figure 4b**); by comparison, just n=4/139 (2.9%) of singleton lineages were ESBL+ (OR 61 [95% CI, 11-422], p<1×10^-7^ for association between ESBL+ and transmission, using Fisher’s exact test). ESBL carriage was a strong predictor of onward transmission of a lineage: we estimate a crude probability of onward nosocomial transmission to be 28% (n=8/29, [95% CI, 11-44%]) for unique ESBL+ strains and 1.7% (n=4/236, [95% CI, 0-3.3%]) for ESBL-strains.

Overall, probable transmission clusters accounted for 55% of all ESBL+ infection episodes and 6.1% of ESBL-infection episodes (**Figure 4c**). Assuming the first clinical isolate from each cluster represents the index case, this implies that 29 infection episodes (9.9%), including 21 ESBL+ infections (45%), resulted from nosocomial transmission (note this is a lower limit as it is possible that the first clinical isolate was also acquired in the hospital from an unsampled source, such as asymptomatic colonization of another patient or staff member, or an environmental reservoir). Transmission-linked ESBL+ infections occurred throughout the study period but were concentrated in Dec 2013 to Feb 2014 (**Figure 4d**), during which time they accounted for 88% of ESBL+ infections (vs 38% in earlier months, p=0.005 using proportion test). This was associated with clusters of ST29 (n=8 patients) and ST323 (n=4 patients) bearing the same *bla*_CTX-M-15_ plasmid (described previously^12^), and the highly resistant ST231 strain noted above (n=3 patients).

#### Common clones not associated with nosocomial transmission

The probable nosocomial transmission clusters account for only seven of the 21 common lineages (ST29, ST45, ST231, ST323, ST340, ST491, ST681). A further three common lineages carried acquired virulence factors (ST86, *ybt* plus virulence plasmid, n=4 patients; ST792, *ybt* plus *clb*, n=4 patients; ST133, *ybt* plus *clb*, n=3 patients) and were associated with community onset (see **Table S3**). The remaining 11 common lineages were fairly typical in terms of AMR (89% susceptible, vs 80% for all other infections, p=0.2), yersiniabactin (34% vs 24%, p=0.1) and onset (53% day 0-2, vs 54%, p=0.9), and it is unclear why they should be detected frequently in clinical infections. Notably, these lineages are also common in clinical isolate collections from other settings (see **Methods**): genomes of ST17, ST20, ST35, ST37, ST111, ST336, ST629, ST661 have all been reported in five other continents (Africa, Asia, Europe, North America and Latin America; n=12-200 public genomes each) and ST27, ST221 and ST412 in two or three continents (n=11-17 public genomes each). In contrast, amongst the other 156 lineages detected in this study, just n=15 (9.6%) have been sequenced from five other continents and n=71 (46%) have not been sequenced from any other continents.

#### K. pneumoniae genomic factors and infection onset

Only 38 infections were classed as true community-onset, and these were not significantly different from healthcare linked infections in terms of age, sex, AMR, hypervirulence determinants, or K/O types (**Table S7a**). Nosocomial onset of infection (i.e., day 3 or later of hospital stay, vs earlier or outpatient onset) was significantly positively associated with male sex, ESBL carriage and *rml*-positive capsule, independently and in a multivariable model (see **Table S7b**). Similar results were observed when including those with a prior hospital admission within 30 days in the definition of nosocomial onset (**Table S7c**).

## Discussion

Here we analysed all clinical isolates identified as *K. pneumoniae* in a hospital clinical microbiology laboratory for a one-year period (**Figure 1, Table 1**), and found remarkable genomic diversity in the underlying population of organisms (**Figures 2, 3**). Consistent with previous studies, we found that 19% of isolates identified as *K. pneumoniae* by MALDI-TOF in fact belonged to other common members of the wider *K. pneumoniae* complex (**Table S2**)^12,22,30,31^. However even amongst the isolates confirmed as *K. pneumoniae*, in this single 1-year local snapshot of disease-associated strains, we detected huge genetic diversity in the form of 138 phylogenetic lineages bearing 78 distinct capsular biosynthesis loci (half of all K loci ever described), 60 acquired AMR genes and 55 plasmid replicons, with just 80% of genes shared pairwise between lineages (**Figures 2 and 3**).

The sheer scale of genomic diversity associated here with clinical disease supports the view of *K. pneumoniae* as a classic opportunistic pathogen, in which any member of the population has the potential to cause disease in hospitalized patients whose underlying health is sufficiently compromised, and that much of the hospital-associated disease burden stems from extraintestinal ‘escape’ by the patients’ own colonizing strains rather than transmission within hospitals^12,31,65^. Indeed only 10% of infections in this study were attributed to WGS-supported nosocomial transmission. Furthermore, our data suggest that most of the common lineages – defined here as the 21 lineages (11.5%) that were detected in ≥3 patients, and which accounted for 38% of total infections – were not transmitted within the hospital system but were detected in multiple patients because they circulate widely in the human population, and in many cases showed evidence of global dissemination. The reasons for the apparent success of these global clones within the human host population are not yet clear; however, as a group they were enriched for ESBL genes, the *ybt* locus, as well as *man*-positive and *rml*-negative capsule types. Notably, many of these lineages (including ST17, ST35, ST37, ST111, ST629, ST661) are amongst those frequently detected in food animals (cows, pigs and/or poultry^66–69^), so may constitute animal-adapted strains to which humans are frequently exposed via the food chain.

Our data reveal a clear association between AMR and nosocomial onset (using either definition). Notably, whilst nosocomial transmission added relatively little to the overall infection burden (∼10%), we estimate that it roughly doubled the burden of ESBL infections. Consistent with this, we estimate the crude risk of transmission resulting in secondary infection/s was negligible for non-ESBL infections (95% CI, 0-3.3%) but substantial (95% CI, 11-44%) for ESBL infections. These observations support the notion that interventions aimed at preventing cross-transmission in hospitals (e.g., hand hygiene, or seek-and-contain approaches to CP infections) could have a significant impact on reducing the total burden of AMR infections. However, the data also suggest that the underlying burden of opportunistic *K. pneumoniae* infection, which originate from diverse strains present in the gut microbiome of patients, might still remain high unless this source of infection is specifically targeted (e.g. by colonization or colonization-density screening)^70^.

One caveat of these analyses is the focus on WGS-supported nosocomial transmission using only simple genetic and temporal distance rules (see **Methods**). However we note our findings are concordant with the detailed epidemiological analyses of probable transmission chains we have reported previously as part of related studies conducted contemporaneously with this one at the Hospital A ICU^31^ and at Hospital C^12^, which incorporated patient movement data and results of carriage screening to detect silent transmission within these at-risk patient subpopulations. The published analyses provided strong evidence for transmission of ST231, CG323, and ST681 in ICU^31^; and for transmission of CG29, CG323 and ST340 more widely in Hospital A, which together could account for all instances of MDR *K. pneumoniae* colonization and infection detected at Hospital C (to which Hospital A geriatric patients are often referred for longer term care)^12^. Similar detailed analyses of ST27, ST35, ST111, ST133, ST412, and ST792 in the Hospital A ICU found no evidence to support intra-hospital transmission of these clones, consistent with the analysis in the present study^31^. Hence, we consider the lack of detailed patient movement data to confirm transmission of the novel clones identified in the present study to be a minor limitation.

Besides AMR, we noted some significant associations between other bacterial factors and infection traits. Virulence plasmid-encoded loci (aerobactin, salmochelin, *rmp*) were associated with known hypervirulent clones and community onset of infection, often with diagnosis made upon presentation to the emergency department or ICU (**Table S3**). The *ybt* locus was associated with common lineages (**Figure 4a**), consistent with previous observations that *ybt* is enriched amongst clinical infection isolates compared to asymptomatic carriage isolates (in the range of >30% vs <10%)^22,24,71^; a recent report that *ybt+* ST258 have a higher attack rate than *ybt*-ST258 in colonised patients^26^; and the known mechanism by which yersiniabactin can enhance the potential for extraintestinal infection, by evading Lcn2-mediated host immunity^25,72^. Notably the presence of the *man* operon in the K locus (correlated with presence of mannose in the expressed capsule^73^) was also associated with common lineages (**Figure 4a**). Mannose-containing capsules have been shown to be recognised by the mannose receptor on human and murine macrophages, promoting clearance and resulting in lower virulence (higher LD_50_) in a murine infection model^74^. Hence, we hypothesise that any advantage conferred by *man*+ K loci likely relates to the process of colonization rather than infection. Consistent with this, the overall prevalence of *man*+ K loci in our clinical isolates (64%) was the same as in a collection of n=464 community carriage isolates recently published from Norway^71^ (65%). Presence of the *rml* operon in the K locus (expected to produce rhamnose-containing capsule^73^) was negatively associated with common lineages (**Figure 4a**) and positively associated with nosocomial onset (using either definition, **Table S7b-c**) but not nosocomial transmission. Indeed, the frequency of *rml*+ was particularly high amongst patients whose first clinical isolate was collected after at least a week into their hospital stay (41% vs 23%; OR 2.3, 95% CI, 1.3-4.2, p=0.003 using Fisher’s exact test). Thus, we hypothesise that *K. pneumoniae* with rhamnose-containing capsules have reduced virulence, as they are apparently less able to establish an infection until the patient’s condition deteriorates sufficiently in hospital. Consistent with this interpretation, the frequency of *rml*+ KL was low (10%) amongst infections diagnosed in the emergency department.

In contrast to the findings above, we did not find any evidence of association between infection site and genomic features of the bacteria, but rather infection site was associated with patient demographics and infection onset: UTIs were significantly associated with age and female sex; respiratory infections with nosocomial onset; and wound infections with community onset. This is consistent with there being no genetically-determined tissue tropism (or limited effect size of bacterial factors, which were underpowered to detect here), and suggests the outcome of the host-pathogen interaction is primarily determined by the patient’s health status and vulnerabilities, consistent with the concept of opportunistic infection. The likely exception is the so-called ‘hypervirulent’ strains that express virulence plasmid-encoded siderophores and hypermucoidy^18,20,21^; such strains were rare in our study, but were mostly detected in community-onset infections (bold in **Table S3**). The spectrum of *K. pneumoniae* disease identified here (∼two-thirds UTI, 15% respiratory, ∼10% wound, ∼10% sepsis) mirrors patterns in other hospitals around the world^75^, and the genomic diversity we uncovered is consistent with WGS studies of unselected bloodstream isolates^18,40,41^ and even from AMR-selected studies^39,75,76^; hence our results are likely to be broadly representative of the *K. pneumoniae* clinical picture in other hospital settings.

Aside from *K. pneumoniae*, the clinical significance of the other species in the *K. pneumoniae* complex remains open to investigation, although it is clear that both *K. variicola* and *K. quasipneumoniae* are capable of causing disease in hospitalized patients^30,77,78^. However, neither of these species was particularly prevalent in our study (18% of all infections, combined). Interestingly, within just a single year of collection in our hospital, we detected 12 *K. variicola* / *K. pneumoniae* hybrid isolates (8 unique strains or variants), all of which involved imports of *K. pneumoniae* into a *K. variicola* background and resulted in import of a capsular biosynthesis locus from *K. pneumoniae* (**Figure S5**). Large-scale recombination between *K. pneumoniae* lineages, centred mainly around the K locus, has been reported several times; the best-known example is the emergence of the carbapenemase clone ST258^79,80^. However to our knowledge very few species hybrids have been reported previously^22,66,81^, and those reported appear to represent sporadic isolations consistent with the expectation that cross-species hybrids have compromised fitness. However, in the present study the KL10 ST681 hybrid strain showed evidence of local transmission in the hospital, spreading to cause silent gut colonization of three patients and various infections in six patients, demonstrating that it is clearly fit to transmit. Notably, the imported capsular synthesis locus KL10 is relatively common in *K. pneumoniae* clinical isolates^54^, with a recent study of the >13,000 publicly available *K. pneumoniae* genomes in GenBank reporting frequencies of 2.1% amongst human blood isolates, 1.5% amongst other human isolates, and 0.3% and 0.5% amongst animal and environmental isolates, respectively^52^. Hence, we hypothesise this may have contributed to the hybrid strain’s ability to colonise or infect humans.

## Conclusions

Overall, we show that by sequencing all clinical isolates, we can gain a much more nuanced view of the burden of *K. pneumoniae* infections. WGS clarified the burden of infection in this setting resulted mainly from diverse strains present in the patients’ own gut microbiomes, including a very low frequency (<3%) of hypervirulent strains and enriched for a small number of successful lineages associated with AMR, yersiniabactin, and *man*+ K loci; on top of which there was an additional burden (∼10%) resulting from nosocomial transmission that is strongly associated with ESBL.

## Data Availability

All data produced in the present work are contained in the manuscript and/or are available online at: https://microreact.org/project/gVB3ki6iA62RREC1stLLHo-kaspah-clinical-isolates.

## Declarations

### Authors’ contributions

Conceptualisation: AWJ, KEH. Methodology and Software: KEH, KLW, MMCL, RG, RRW. Formal Analysis and Visualisation: CLG, KEH, KLW, MMCL, RG, RRW. Investigation: DWS, DVP, IJA, LMJ, MM, NFP, PCH, SAM. Resources: NRT, RAS. Data curation: JSG, KMW. Supervision: AWJ, KEH, RAS. Funding: AWJ, KEH. Writing – Original Draft Preparation: CLG, KEH, KLW. All authors read and approved the final manuscript.

## Acknowledgments

The authors gratefully acknowledge the contribution and support of Janine Roney, Mellissa Bryant, Jennifer Williams and Noelene Browne at the Alfred Hospital; Elke van Gorp of Monash University for re-testing fosfomycin susceptibility of selected isolates; and the sequencing team at the Wellcome Sanger Institute. We also thank the team of curators at the Institut Pasteur MLST system (Paris, France) for importing novel alleles, profiles and/or isolates at bigsdb.web.pasteur.fr.

## Data availability

Raw Illumina reads are available in ENA, accessions for each genome are in **Table S1**. Reference-quality hybrid Illumina+ONT assemblies are available in GenBank, accessions are included in **Table S1**. Illumina-only assemblies were not deposited as they are considered draft assemblies, however the full set of assemblies and pan-genome data used in this study are available in FigShare (doi: 10.26180/16811344). The annotated phylogeny is available via MicroReact (microreact.org/project/gVB3ki6iA62RREC1stLLHo-kaspah-clinical-isolates).

