## Supplementary Figures and Tables for "Genomic dissection of the bacterial population underlying *Klebsiella pneumoniae* infections in hospital patients: insights into an opportunistic pathogen"

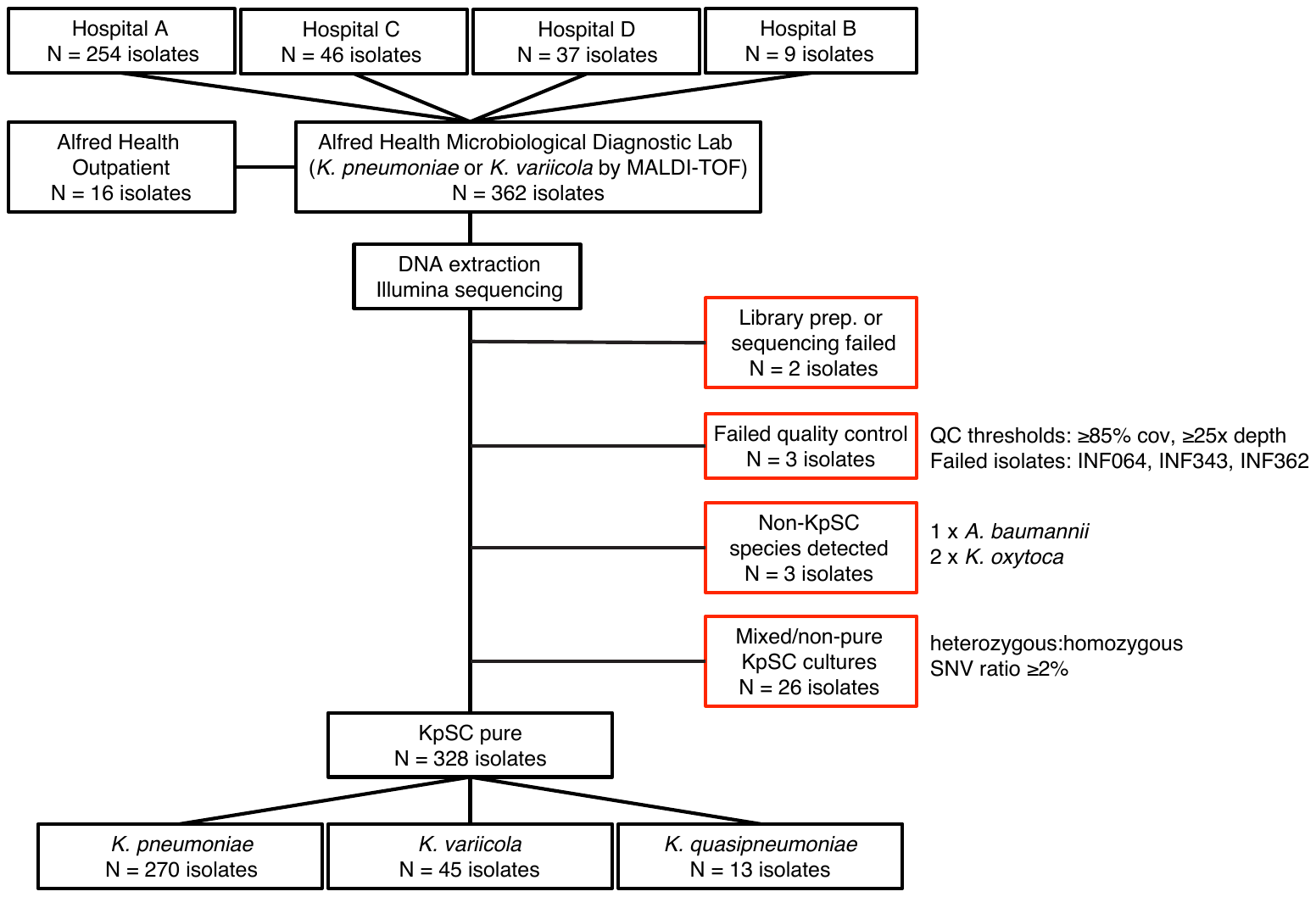

**Figure S1. Flowchart for collection and sequencing of clinical isolates.**

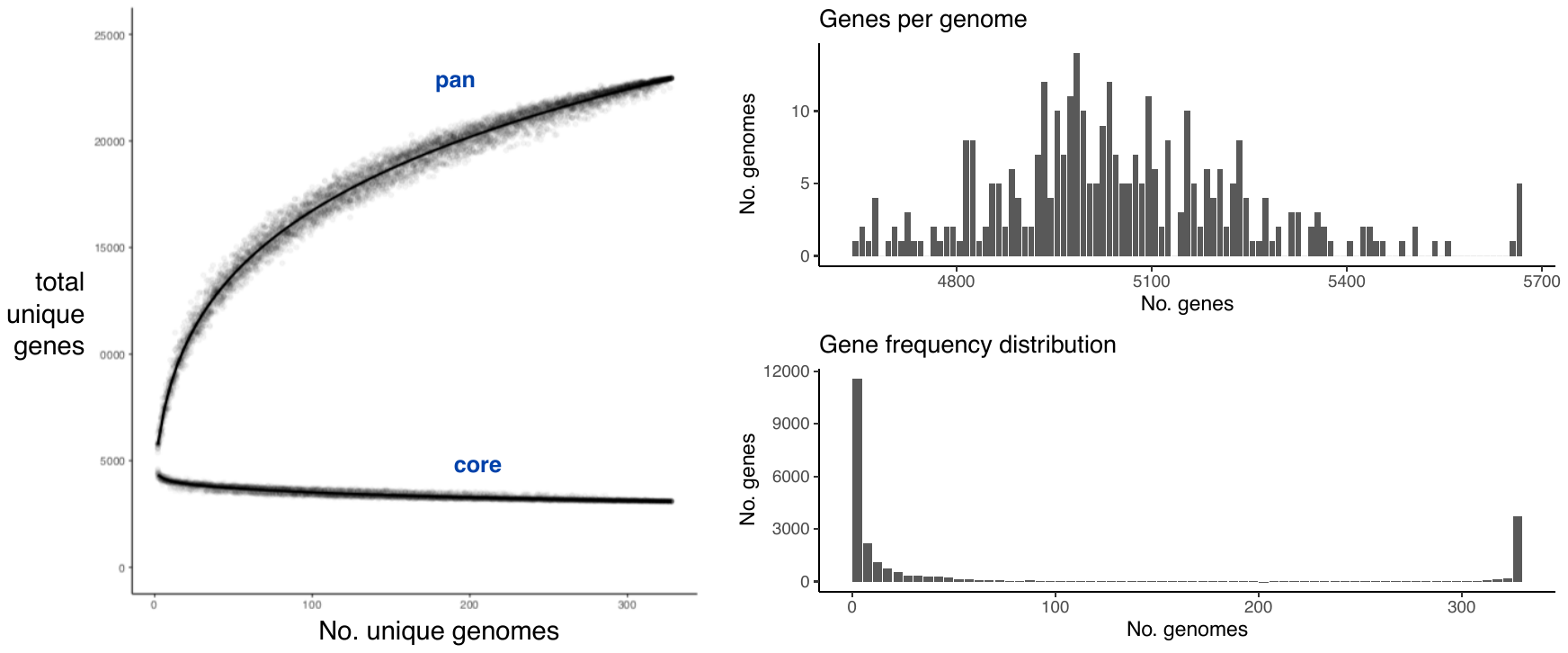

**Figure S2. Pan genome.** Data used for this analysis was all KpSC genomes.

(a) Upper curve shows the pan genome, i.e. total unique genes encountered (y-axis) in a sample of *n* genomes (x-axis). Lower curve shows the core genome, i.e. total genes shared by all genomes (y-axis), in a sample of *n* genomes (x-axis). Points correspond to 20 random samples of *n* genomes, for *n*=1, …, 328; curves are Loess smoothed curves plotted through these points using the geom_smooth() function in R, with the formula ‘y~log(x)’. (b) Distribution of total number of genes per genome. (c) Distribution of gene frequencies across the set of 328 genomes.

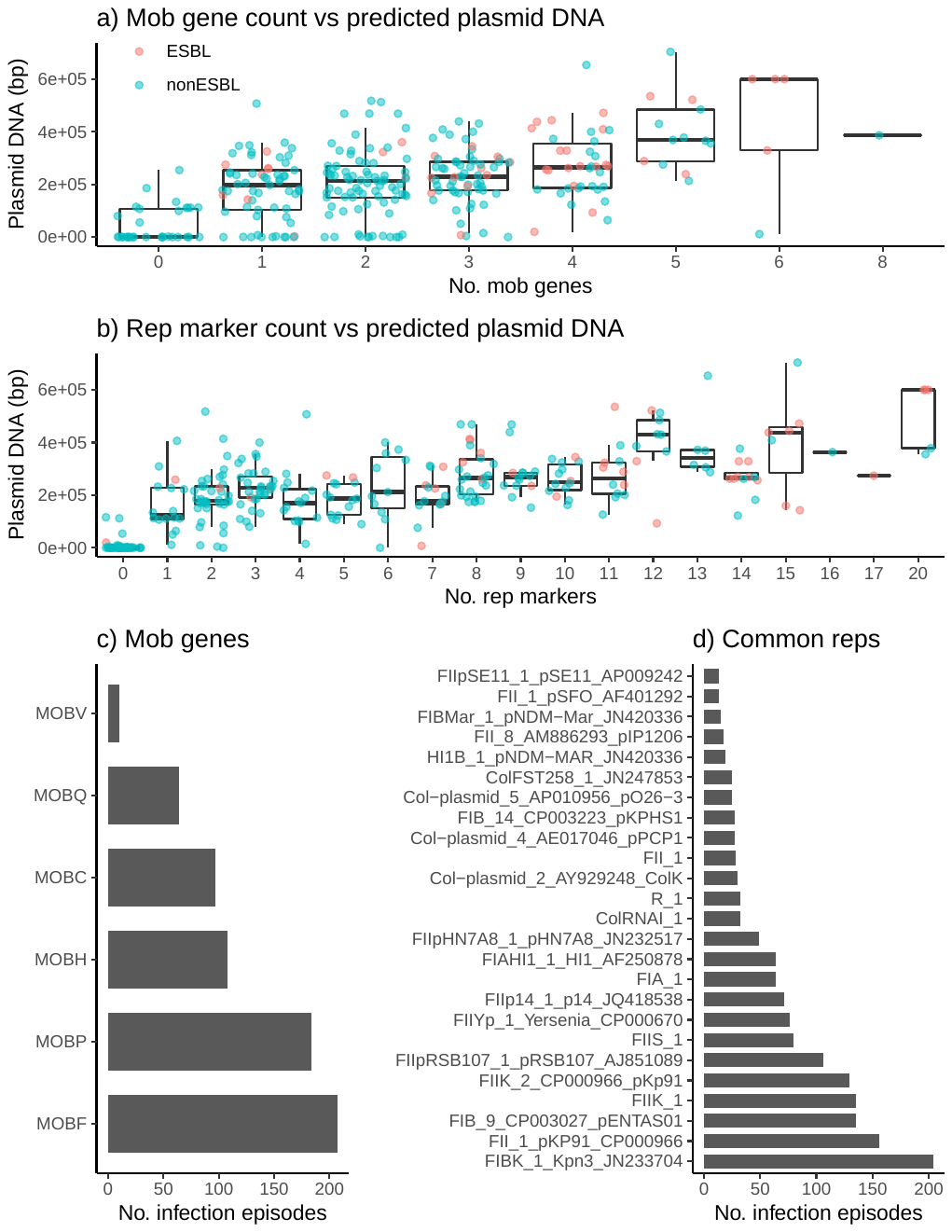

**Figure S3. Measures of plasmid number and diversity.** Data used for this analysis was the first isolate per unique infection episode, for 294 genomically-defined infection episodes. Distribution of total DNA sequence (bp) in contigs assigned as plasmids are shown, stratified by (a) number of *mob* genes identified per genome and (b) number of uniquely distributed replicon markers identified per genome. Each point represents a unique genome sequence and is coloured according to whether ESBL genes were detected in the genome (inset legend). Linear regression model fit: Total plasmid DNA (kbp) ~ 79 + 18 * (#*mob*) + 16 * (#*rep*); p=8x10^-4^ for *mob*; p<1x10^-15^ for *rep*. (c) Frequency of individual *mob* types. (d) Frequency of 25 common replicon markers (each present in ≥5% of infections).

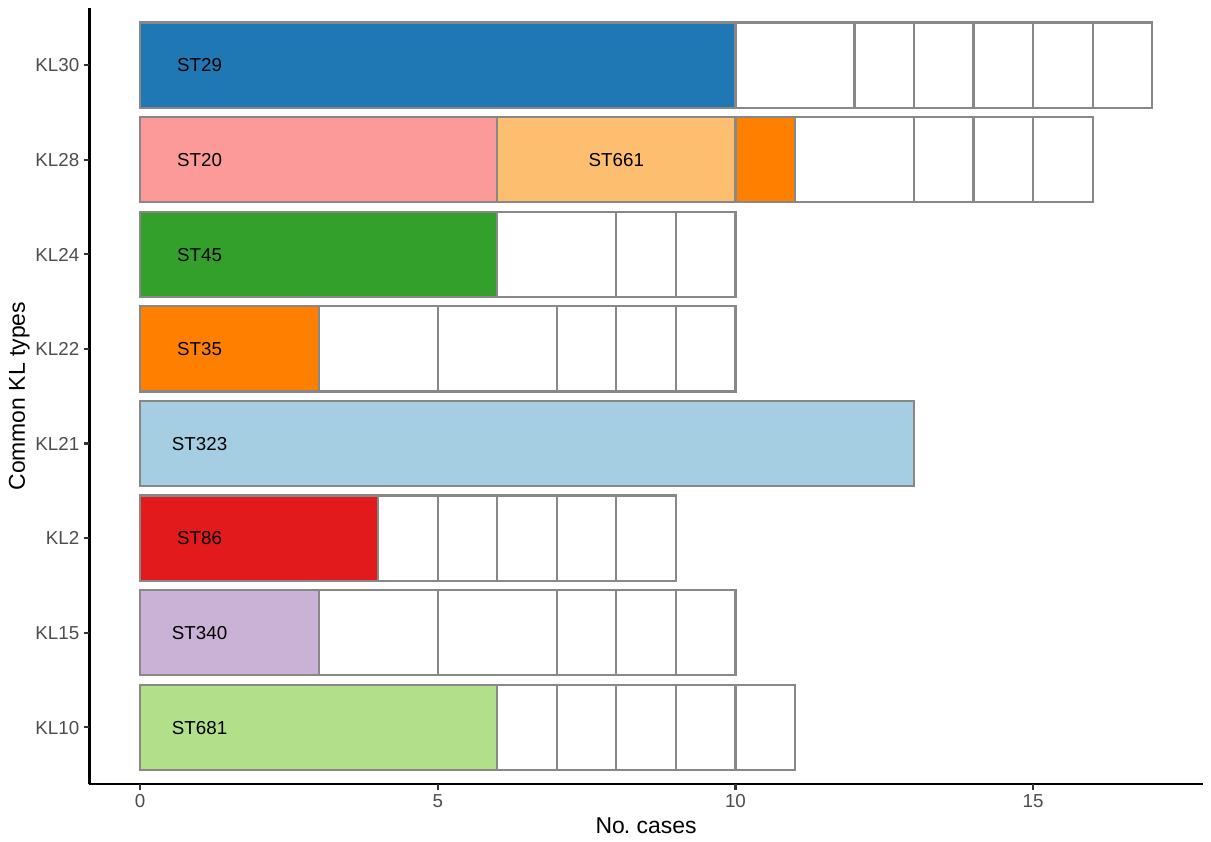

**Figure S4. Eight most common K locus types.** Data used for this analysis was the first isolate per unique infection episode, for 294 infection episodes. Stacked bars represent different K locus (KL) types and are stratified by ST; common STs are coloured and labelled; rare STs are coloured white.

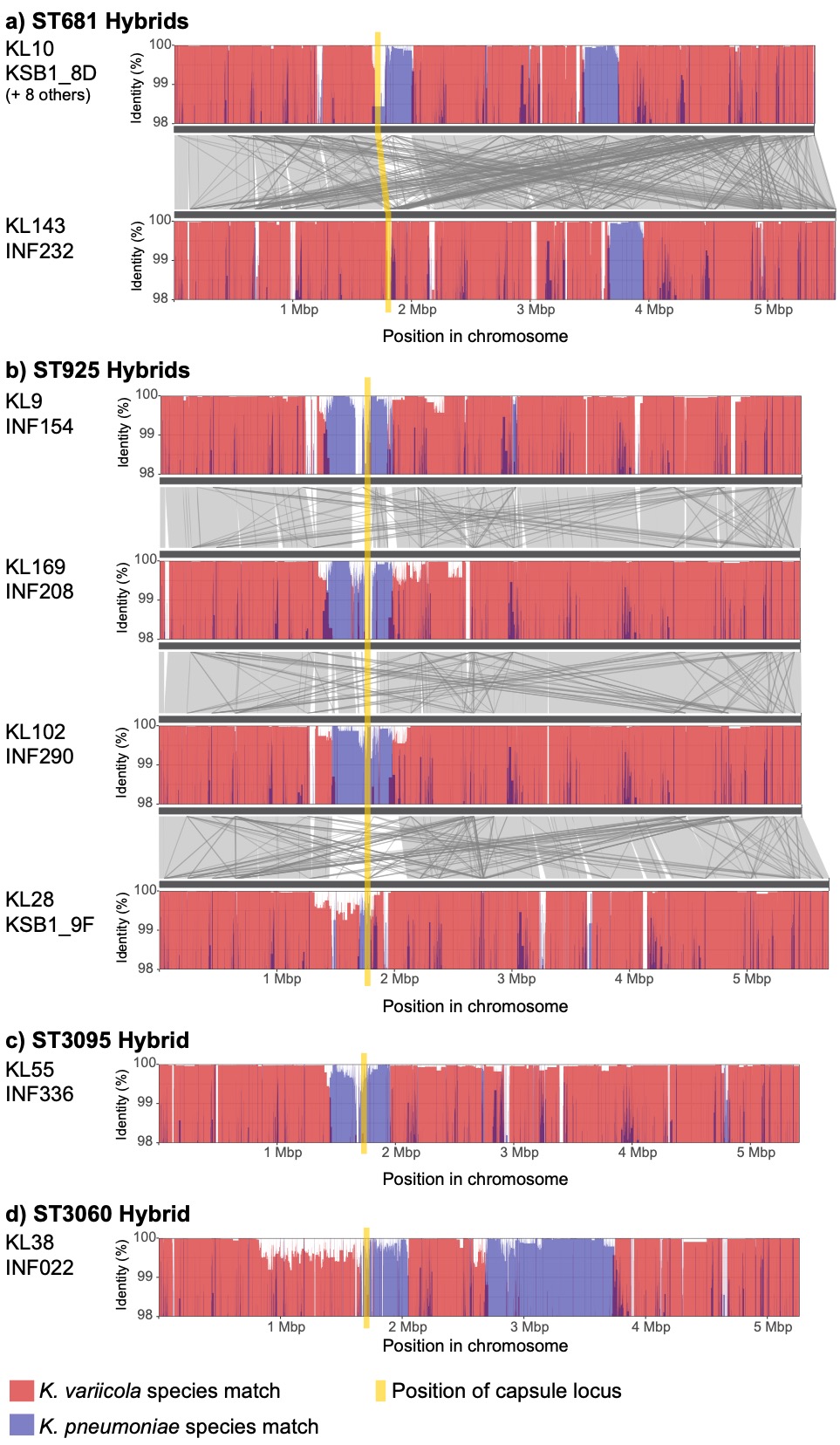

**Figure S5. Details of hybrids identified between KpSC species.** Plots show nucleotide identity to *K. variicola* (red) or *K. pneumoniae* (blue) sequences in sliding windows along each genome. For (a) and (b), homology between different genomes is shown in grey. Position of the capsule (K) biosynthesis locus is shown in yellow and is directly adjacent to the outer lipopolysaccharide (O) synthesis locus.

|  | ***K. pneumoniae*** | ***K. variicola*** | ***K. quasipneumoniae*** |
| --- | --- | --- | --- |
| **Any acquired AMR (p=0.02)** | 79 (33%) | 7 (16%) | 1 (8%) |
| **ESBL+ (p=0.3)** | 42 (18%) | 4 (9%) | 0 |
| **MDR (p=0.07)** | 49 (21%) | 3 (7%) | 1 (8%) |
| **Ybt+ (p=5x10^-6^)** | 78 (33%) | 0 | 0 |
| **Iuc+ (p=0.39)** | 8 (3%) | 0 | 0 |
| **Infection Onset (p=0.74)** |  |  |  |
| Day 0-2 | 129 (54%) | 22 (50%) | 8 (67%) |
| Day 3+ | 97 (41%) | 21 (48%) | 4 (33%) |
| Outpatient | 12 (5%) | 1 (2%) | 0 |
| **Acquisition (p=0.47)** |  |  |  |
| Nosocomial | 120 (50%) | 27 (61%) | 4 (33%) |
| Healthcare | 87 37%) | 12 (27%) | 6 (50%) |
| Community | 31 (13%) | 5 (11%) | 2 (17%) |
| **Infection site** |  |  |  |
| Urinary tract | 161 (68%) | 25 (57%) | 7 (68%) |
| Respiratory | 33 (14%) | 7 (16%) | 3 (14%) |
| Disseminated | 18 (8%) | 6 (14%) | 1 (8%) |
| Wound/Tissue | 23 (19%) | 4 (9%) | 1 (10%) |
| Other | 3 (1%) | 2 (5%) | 0 (1%) |
| **N (% of total)** | 238 (81%) | 44 (15%) | 12 (4%) |

**Table S2. Frequency of genetic features and infection characteristics, by species.** Data shown represents one sequenced isolate per unique infection episode (n=294).

Cells show number of isolates of each species (column) that exhibit a given trait (row).

Percentages in brackets indicate the frequency of the trait within the given species; except for the last row which shows the total number of infection episodes for each species, and the percentage this represents of the total infections. ESBL = extended spectrum beta-lactamase; MDR = resistance to ≥3 drug classes except ampicillin; Ybt = yersiniabactin, Iuc = aerobactin.

| **Isolate** | **Age range** | **Sex** | **Specimen** | **Day**  **(Ward)** | **Lineage** | **ICE*Kp*** | **Vir plasmid** | **K** | **O** |
| --- | --- | --- | --- | --- | --- | --- | --- | --- | --- |
| INF191 | 60-69 | M | Respiratory | 1 | **ST86** | *ybt9* | *iuc1, iro1, rmp1* | KL2 | O1 |
| INF225 | 60-69 | F | UTI | 1 (ED) | **ST86** | *ybt9* | *iuc1, iro1, rmp1* | KL2 | O1 |
| INF254 | 80-89 | F | UTI | (OP) | **ST86** | *ybt9* | *iuc1, iro1, rmp1* | KL2 | O1 |
| INF331 | 80-89 | F | Disseminated | 1 (ED) | **ST86** | *ybt9* | *iro1, rmp1* | KL2 | O1 |
| INF079 | 80-89 | F | UTI | 0 (ED) | **ST23** | *ybt1, clb2* | *iuc1, iro1, rmp1* | KL1 | O1 |
| INF196 | 40-49 | M | Wound/tissue | 0 | **ST66** | *ybt12, clb1* | *iuc2, iro2, rmp2* | KL2 | O1 |
| INF351 | 60-69 | M | Respiratory | 0 | **ST3051^** | *ybt9* | *iuc1, iro1, rmp1* | KL20 | O1 |
| INF237 | 60-69 | F | Respiratory | (OP) | **ST91^§^** | *ybt10* | *iuc2A* | KL4# | O2ac |
| INF151 | 80-89 | M | Respiratory | 18 | **ST93^§^** | *ybt10* | *iuc2A, *rmp2A* | KL4 | O2ac |
| INF078 | 60-69 | F | UTI | 1 | ST105 | *ybt15, *iro4* | *-* | KL102 | O2afg |
| INF038 | 30-39 | F | Disseminated | 1 (ED) | ST60 | *ybt2, iro3, *rmp3* | *-* | KL5 | O1 |
| INF269 | 80-89 | M | UTI | 30 | ST13 | *ybt17, clb3* | *-* | KL3 | O1 |
| INF002 | 40-49 | M | Respiratory | 2 (ICU) | ST133 | *ybt17, clb3* | *-* | KL116 | O1 |
| INF101 | 50-59 | F | UTI | 1 | ST133 | *ybt17, clb3* | *-* | KL116 | O1 |
| INF339 | 20-29 | M | Wound/tissue | 0 (ICU) | ST792 | *ybt17, clb3* | *-* | KL25# | O3/O3a |
| INF227 | 20-29 | F | UTI | 0 (ED) | ST792 | *ybt17, clb3* | *-* | KL25 | O3/O3a |
| INF317 | 60-69 | M | Respiratory | 9 (ICU) | ST792 | *ybt17, clb3* | *-* | KL25 | O3/O3a |
| INF341 | 50-59 | F | UTI | 0 (ED) | ST792 | *ybt17, clb3* | *-* | KL25 | O3/O3a |

**Table S3. Infections associated with carriage of acquired virulence factors.** Bolded sequence types (STs) indicate known hypervirulent lineages commonly associated with carriage of both ICE*Kp* and the virulence plasmid. Day=day of onset of infection, relative to current hospital admission, Ward indicates the location of the patient at the time the specimen was taken for culturing: OP=outpatient, ED=emergency department, ICU=intensive care unit. K=capsule biosynthesis locus, O=predicted O (lipopolysaccharide) antigen. ^Single-locus variant of the virulence plasmid-associated ST420. ^§^*K. pneumoniae* subsp. *ozaenae*. *Locus incomplete, most likely not functional. #Low confidence K locus match. Note a third infection with ST133, but lacking *ybt* and *clb*, was also identified (INF072, male in his 70s who developed pneumonia on day 4 of ICU stay).

| **KL** | **No. infections** | **No. STs** | ***man* operon** | ***rml* operon** |
| --- | --- | --- | --- | --- |
| KL30 | 17 | 7 | yes | no |
| KL28 | 16 | 7 | yes | no |
| KL21 | 13 | 1 | yes | no |
| KL10 | 11 | 6 | yes | no |
| KL15 | 10 | 6 | no | no |
| KL22 | 10 | 6 | no | no |
| KL24 | 10 | 4 | yes | no |
| KL2 | 9 | 6 | yes | no |
| KL14 | 8 | 4 | yes | yes |
| KL25 | 8 | 5 | no | no |
| KL63 | 8 | 5 | yes* | no |
| KL39 | 6 | 2 | yes | no |
| KL47 | 6 | 6 | no | yes |
| KL54 | 6 | 5 | yes* | no |
| KL55 | 6 | 5 | no | yes |
| KL64 | 6 | 4 | yes | yes |
| KL102 | 5 | 3 | no | no |
| KL142 | 5 | 5 | no | yes |
| KL16 | 5 | 4 | yes* | no |
| KL118 | 4 | 1 | no | yes |
| KL38 | 4 | 4 | no | no |
| KL53 | 4 | 3 | yes | yes |
| KL57 | 4 | 3 | yes | no |
| KL116 | 3 | 1 | yes | no |
| KL13 | 3 | 3 | yes | no |
| KL20 | 3 | 3 | yes | no |
| KL3 | 3 | 3 | yes | no |
| KL52 | 3 | 2 | no | yes |
| KL60 | 3 | 3 | yes | no |
| KL7 | 3 | 2 | yes | no |
| KL9 | 3 | 3 | no | yes |
| KL103 | 2 | 2 | no | yes |
| KL105 | 2 | 2 | no | yes |
| KL109 | 2 | 1 | yes | yes |
| KL114 | 2 | 2 | yes | no |
| KL122 | 2 | 2 | yes | no |
| KL124 | 2 | 1 | no | yes |
| KL125 | 2 | 2 | no | no |
| KL143 | 2 | 2 | yes | no |
| KL169 | 2 | 2 | yes | yes |
| KL170 | 2 | 1 | no | yes |
| KL36 | 2 | 2 | no | yes |
| KL46 | 2 | 2 | yes | no |
| KL48 | 2 | 2 | no | yes |
| KL51 | 2 | 2 | no | no |
| KL58 | 2 | 2 | yes* | no |
| KL62 | 2 | 2 | yes | no |
| KL71 | 2 | 2 | no | yes |
| **KL** | **No. infections** | **No. STs** | ***man* operon** | ***rml* operon** |
| KL8 | 2 | 2 | no | no |
| KL1 | 1 | 1 | yes* | no |
| KL101 | 1 | 1 | no | yes |
| KL107 | 1 | 1 | no | yes |
| KL108 | 1 | 1 | yes | no |
| KL11 | 1 | 1 | no | no |
| KL111 | 1 | 1 | no | no |
| KL112 | 1 | 1 | yes | no |
| KL113 | 1 | 1 | yes | no |
| KL119 | 1 | 1 | yes | no |
| KL12 | 1 | 1 | no | yes |
| KL120 | 1 | 1 | no | yes |
| KL123 | 1 | 1 | no | no |
| KL127 | 1 | 1 | no | yes |
| KL130 | 1 | 1 | yes | yes |
| KL131 | 1 | 1 | no | yes |
| KL132 | 1 | 1 | yes | no |
| KL133 | 1 | 1 | yes | no |
| KL134 | 1 | 1 | no | no |
| KL137 | 1 | 1 | yes | yes |
| KL139 | 1 | 1 | yes | no |
| KL140 | 1 | 1 | yes | no |
| KL141 | 1 | 1 | yes | yes |
| KL144 | 1 | 1 | yes | yes |
| KL146 | 1 | 1 | yes | no |
| KL153 | 1 | 1 | yes | no |
| KL158 | 1 | 1 | no | yes |
| KL166 | 1 | 1 | yes | yes |
| KL167 | 1 | 1 | yes | no |
| KL168 | 1 | 1 | yes | yes |
| KL17 | 1 | 1 | no | yes |
| KL18 | 1 | 1 | no | yes |
| KL19 | 1 | 1 | no | yes |
| KL23 | 1 | 1 | no | yes |
| KL34 | 1 | 1 | no | yes |
| KL35 | 1 | 1 | yes | no |
| KL37 | 1 | 1 | no | no |
| KL4 | 1 | 1 | no^ | no |
| KL42 | 1 | 1 | yes | no |
| KL45 | 1 | 1 | no | yes |
| KL5 | 1 | 1 | yes | no |
| KL61 | 1 | 1 | yes | no |
| KL67 | 1 | 1 | yes | yes |
| unknown | 11 | 11 | - | - |

**Table S4. Capsule (K) biosynthesis loci identified in clinical isolates in this study.**

‘No. STs’ indicates the number of unique multi-locus sequence types (STs) each K locus was identified in. Presence of the mannose (*man*) or rhamnose (*rml*) operons are indicated. For KL with known structures (KL1-KL82): presence of *rml* is perfectly correlated with presence of rhamnose in the expressed capsular polysaccharide (CPS); * indicates KL is *man*+ but CPS lacks mannose; ^ indicates KL is *man*- but CPS contains mannose. Sugar structures for CPS encoded by KL100-KL170 are unknown.

| **Species** | **ST** | **No. Patients** | **AMR** | ***ybt*** | **K locus** | **O type** |
| --- | --- | --- | --- | --- | --- | --- |
| *Kp* | ST29 | 9 | ESBL, MDR | - | KL30^m^ | O1 |
| *Kv* | ST681 | 6 | susceptible* | - | KL10^m^ | O2afg |
| *Kp* | ST323 | 4 | ESBL, MDR | - | KL21^m^ | O3b |
| *Kp* | ST323 | 3 | ESBL, MDR | - | KL21^m^ | O3b |
| *Kp* | ST231 | 3 | CP, ESBL, MDR | *ybt* 15 | KL64^m,r^ | O1 |
| *Kp* | ST491 | 3 | ESBL, MDR | - | KL118^r^ | OL101 |
| *Kp* | ST340 | 3 | ESBL, MDR | *ybt* 16 | KL15 | O4 |
| *Kp* | ST2370 | 2 | ESBL, MDR | *ybt* 4 | KL15 | O4 |
| *Kqs* | ST5872 | 2 | ESBL, MDR | - | KL7^m^ | O12 |
| *Kp* | ST45 | 2 | susceptible | *ybt* 10 | KL24^m^ | O2a |
| *Kqs* | ST1548 | 2 | susceptible | - | KL170 | O3/O3a |
| *Kqs* | ST480 | 2 | susceptible | - | KL52^r^ | OL103 |

**Table S5.** Features of probable nosocomial transmission clusters. *Kp, K. pneumoniae; Kv, K. variicola; Kqs, K. quasipneumoniae subsp. similipneumoniae*. ST, sequence type. Patients, number of patients involved in the putative cluster. AMR, indicates presence of antimicrobial resistance including CP (carbapenemase producing), ESBL (extended spectrum beta-lactamase producing), MDR (multidrug resistant, i.e. resistant to ≥3 drug classes in addition to ampicillin). *The last ST681 isolated was ESBL and MDR. *Ybt* column indicates presence of yersiniabactin lineages; K locus column indicates the capsule biosynthesis locus present; O type column indicates the O (lipopolysaccharide) antigen predicted from genome data.

|  | **Transmission** | | |
| --- | --- | --- | --- |
| *Predictors* | *Odds Ratio* | *95% CI* | *p* |
| ESBL+ | **21.0** | **9.21 – 51.1** | **<1x10^-11^** |
| Yersiniabactin | 1.00 | 0.38 – 2.49 | 0.997 |
| Patient Age (years) | **0.97** | **0.95 – 1.00** | **0.017** |
| Patient Sex (male) | 0.89 | 0.37 – 2.07 | 0.779 |
| Onset day ≥3 | **2.64** | **1.12 – 6.42** | **0.028** |
| *man*+ KL | 0.52 | 0.17 – 1.46 | 0.230 |
| *man*+ OL | 1.22 | 0.49 – 3.17 | 0.673 |

**Table S6. Logistic regression model for transmission.**

Results shown are for a multivariable logistic regression model of 294 unique infections, with all variables included as predictors (95% CI = 95% confidence intervals for odds ratios). All variables were coded as binary except for age, which is continuous and expressed in years.

|  | (a) **Community Acquired** | | |
| --- | --- | --- | --- |
| *Predictors* | *Odds Ratio* | *95% CI* | *p* |
| Species *K. pneumoniae* | 0.83 | 0.31 – 2.40 | 0.712 |
| ESBL+ | 0.55 | 0.15 – 1.50 | 0.287 |
| Ybt+ | 2.03 | 0.89 – 4.57 | 0.088 |
| Iuc+ | 0.58 | 0.03 – 3.78 | 0.626 |
| Patient Age (years) | 1.00 | 0.98 – 1.02 | 0.930 |
| Patient Sex (male) | 0.72 | 0.34 – 1.47 | 0.371 |
| *rml*+ KL | 0.67 | 0.26 – 1.58 | 0.378 |
| *man*+ KL | 1.27 | 0.60 – 2.78 | 0.545 |
| *man*+ OL | 1.53 | 0.69 – 3.32 | 0.284 |

|  | **(b) Onset day 3+** | | | **(c) Onset day 3+ and/or**  **recent inpatient admission** | | | |
| --- | --- | --- | --- | --- | --- | --- | --- |
| *Predictors* | *Odds Ratio* | *95% CI* | *p* | | *Odds Ratio* | *95% CI* | *p* |
| Species *K. pneumoniae* | 1.39 | 0.69 – 2.86 | 0.365 | | 1.04 | 0.52 – 2.06 | 0.917 |
| ESBL+ | **2.34** | **1.18 – 4.72** | **0.015** | | **2.03** | **1.04 – 4.10** | **0.042** |
| Ybt+ | 0.79 | 0.42 – 1.47 | 0.465 | | 0.77 | 0.42 – 1.38 | 0.379 |
| Iuc+ | 0.31 | 0.02 – 2.00 | 0.299 | | 0.43 | 0.06 – 2.10 | 0.336 |
| Patient Age (years) | 1.01 | 0.99 – 1.02 | 0.406 | | 1.00 | 0.99 – 1.01 | 0.780 |
| Patient Sex (male) | **2.20** | **1.32 – 3.71** | **0.003** | | **1.72** | **1.05 – 2.82** | **0.031** |
| *rml*+ KL | **3.12** | **1.72 – 5.74** | **<0.001** | | **2.05** | **1.14 – 3.75** | **0.017** |
| *man*+ KL | 0.78 | 0.45 – 1.35 | 0.369 | | 0.77 | 0.46 – 1.30 | 0.332 |
| *man*+ OL | 0.87 | 0.49 – 1.52 | 0.620 | | 0.67 | 0.39 – 1.15 | 0.146 |

**Table S7. Logistic regression models for infection acquisition.**

Results shown are for a multivariable logistic regression model of 294 unique infections, with all variables included as predictors (95% CI = 95% confidence intervals for odds ratios). Predictors with *P*-values below 0.05 are bolded. All variables were coded as binary except for age, which is continuous and expressed in years. (a) Outcome variable = community associated (CA) infection, i.e. isolation from an outpatient or on day 0–2 of current admission as an inpatient, and with no recorded prior contact with the Alfred Health Network (either as an inpatient or outpatient) in the previous 12 months (n=38 CA vs 256 non-CA). (b) Outcome variable = nosocomial onset, defined as isolation on day 3 or later of the current inpatient admission (n=122 vs 172). (c) Outcome variable = nosocomial, defined as isolation on day 3 or later of the current inpatient admission or with recent inpatient admission (in the last month) (n=151 vs 143).
